# Brainstem functional connectivity integrates microbiome-gut-brain signals in healthy older adults: a cross-sectional evaluation and a randomised controlled probiotic trial

**DOI:** 10.64898/2026.11.04.26366206

**Authors:** Julia Rode, Ashley N Hutchinson, Per Thunberg, Robert J Brummer

**Affiliations:** School of Medical Sciences, Faculty of Medicine and Health, Örebro University, 70182 Örebro, Sweden; Food and Health Centre, Örebro University, 70182 Örebro, Sweden; Department of Radiology and Medical Physics, Faculty of Medicine and Health, Örebro University, 70182 Örebro, Sweden; Center for Experimental and Biomedical Imaging in Örebro (CEBIO), Faculty of Medicine and Health, Örebro University, 70182 Örebro, Sweden

**Author notes:** Corresponding author: Julia Rode, 0046 19 30 3000, Örebro University, Fakultetsgatan 1, 70182 Örebro, Sweden, Highest academic titles: Julia Rode PhD, Ashley N Hutchinson PhD, Per Thunberg Associate Professor, Robert J Brummer Professor.

**Keywords:** brainstem, resting state, functional connectivity, functional magnetic resonance imaging, fMRI, ageing, probiotic, gut-brain

## Abstract

**Background:** Despite increasing interest in the gut-brain axis, the role of the autonomic nervous system and the brainstem in human gut-brain signalling remain largely unknown.

**Objective:** To characterise resting state functional connectivity (FC) of the brainstem in healthy older adults and to examine its association with a variety of factors relevant to ageing and the gut-brain axis, and to assess if brainstem FC is altered by probiotic intervention.

**Design:** Randomised, double-blinded, placebo-controlled trial in 60–80-year-olds (55 women/28 men) with six-weeks *Lacticaseibacillus rhamnosus* HN001 intervention in a non-encapsulated and microencapsulated formulation. Brainstem-optimised functional magnetic resonance imaging was performed before and after intervention.

**Results:** At baseline, age correlated positively with FC of locus coeruleus (LC) and dorsal raphe nucleus (DRN) with pre-/postcentral gyrus, as well as of periaqueductal gray (PAG) with temporal/supramarginal gyrus and planum temporale. FC of PAG with precuneus cortex and cingulate gyrus correlated with cognitive status and mood. Perceived stress correlated with FC between nucleus tractus solitarius and occipital cortex, angular/temporal gyrus. FC of DRN differed between men and women. All p<0.0063.

Following probiotic intervention, a significant time*group interaction and main time effect were observed for FC of LC and ventral tegmental area. FC of both with occipital pole/cortex and cerebellum, and of LC with pre-/postcentral gyrus, superior parietal lobule, insular/opercular cortex, putamen, and planum polare showed within-group changes (non-encapsulated), no between-group differences were observed (all p<0.0056).

**Conclusion:** Brainstem-to-cortex FC at rest correlates with several phenotypic characteristics implicated in healthy ageing and may be modifiable by probiotic supplementation.

**KEY MESSAGES:** What is already known on this topic: Gut-derived signals, including those related to gut microbiota modulations, influence brain function. Whether and how those signals are transmitted via the autonomic nervous system and integrated via the brainstem remains largely unexplored in humans.

What this study adds: Brainstem-to-cortex functional connectivity, originating especially from the periaqueductal gray and locus coeruleus, was associated with age, cognitive status and psychological symptoms; and functional connectivity especially of the locus coeruleus was also affected by probiotic intervention.

How this study might affect research, practice or policy: From a human gut-brain perspective, imaging the brainstem may add to the understanding of the role of the autonomic nervous system in (patho-)physiological processes and modes-of-action of intervention effects. This may have clinical implications for conditions with disturbed signalling along this axis.

## INTRODUCTION

The brainstem plays a key role in integrating peripheral physiological signals^1^. This is especially interesting when considering the gut-brain axis, as gut-derived signals along the enteric and autonomic nervous systems, including those associated with dietary compounds or gut microbiota modulations, are integrated within the brainstem^1^. While research on the gut-brain axis has been growing extensively, the role of the brainstem in mediating communication along this axis remains relatively unexplored^2^.

Functional brain imaging has predominantly focused on evaluations of the cortex. However, advancements in neuroimaging techniques, including functional magnetic resonance imaging (fMRI), have made concurrent investigations of brainstem and cerebrum in humans more feasible.

Recent studies typically assessed resting state brainstem functional connectivity (FC) in healthy young to middle-aged cohorts in cross-sectional studies. In these populations, brainstem-to-cortex FC has been shown to correlate with age^3^, depression symptom scores^4^, cognitive performance^5^ and working memory^6^, as well as to differ by sex^3^. Intriguingly, brainstem FC of older adults (55–80 years of age) has been reported to differ from 20–40-year-old adults^6^. This is especially relevant, because brainstem regions have been identified as connector hubs linking multiple brain networks, including default mode and executive control^7^, which both are known to undergo ageing-related alterations^8^.

Very few studies have assessed brainstem FC at more than one timepoint, and those that have typically focused on short-term changes following acute challenges.

In terms of gastrointestinal tract-to-brainstem signalling and activation of brainstem nuclei, gastric stimuli received the most attention so far. Studies assessing FC of the brainstem have done so in relation to gastric motility^9^, stomach-brain coupling^10^, acute stimulation through duodenal capsaicin^11^ and oral intake of low or high-fat yoghurt^12^ or sucrose^13^. Consistent with its involvement in gut-brain signalling, altered brainstem-cortex resting state FC has been reported in individuals with irritable bowel syndrome^14^ ^15^.

Intervention studies specifically assessing brainstem FC, to the best of our knowledge, have not yet been conducted. A well-controlled, gut-targeted intervention that has been shown to affect brain function is probiotic supplementation^16^. Prolonged (four to six weeks) regular intake of probiotics affected (cortical) resting state FC in healthy young/middle-aged populations^17–19^ as well as in elderly^20^. Tillisch et al. even reported altered FC of a network including the midbrain during resting state and an emotional attention task^19^. However, these studies examined broad patterns of brain FC rather than specifically examining brainstem FC.

Thus, the herein presented study investigated two underexplored aims: i) to characterise baseline FC of the brainstem in healthy older adults and to determine association with a variety of factors relevant to ageing and the gut-brain axis, such as age, sex, cognition, mental health, stress perception, sleep, gastrointestinal symptoms, and inflammation; ii) to determine if resting state brainstem FC is altered by a probiotic intervention which consisted of six-weeks daily supplementation with *Lacticaseibacillus rhamnosus* HN001.

## METHODS

Brainstem-optimised functional magnetic resonance imaging (fMRI) examinations were performed in an elderly community-dwelling population before and after a six-weeks randomised, double-blinded, placebo-controlled daily probiotic intervention study with three parallel arms: micro-encapsulated or non-encapsulated *Lacticaseibacillus rhamnosus* HN001 or placebo.

### Intervention study

Participants of both sexes were recruited through local advertisement, screened at the study site and included if 60–80 years of age, body mass index 18.5–31.9 kg/m^2^, willing to abstain from regular consumption of probiotic supplements or food products containing probiotic bacteria (including fermented food/beverages) and supplements/medications known to alter gastrointestinal function or inflammatory status, and self-reported healthy (e.g. lacking a metabolic, gastrointestinal, psychiatric, neurodegenerative diagnosis). Participants were phenotypically characterised (and intervention effects were assessed^20^) by researcher-administered tests (Montreal Cognitive Assessment (MoCA)^21^), self-administered questionnaires assessing mental health and sleep quality (Hospital Anxiety and Depression Scale (HADS)^22^, Perceived Stress Scale (PSS)^23^, Pittsburgh Sleep Quality Index (PSQI)^24^, Gastrointestinal Symptom Rating Scale (GSRS)^25^) and blood markers (high sensitivity C-reactive protein (hsCRP), Supplementary Information 1). Block (size three and six) randomisation was performed, and participants were stratified by sex and age (born 1954 or later, born 1953 or earlier). Randomisation and allocation (ratio 1:1:1) were performed by colleagues not involved in the study, resulting in a sequentially numbered list. The study was blinded, and allocation sequence was inaccessible (digitally stored) to participants, caregivers, researchers and analysts until primary and some secondary/exploratory outcome analyses were complete. The analyses presented herein were performed unblinded as this was an exploratory analysis of the original study. For the same reason, the concrete statistical analyses had not been predefined. *A priori* sample size calculation was based on the primary outcome, *i.e.* intervention effects on cortical functional connectivity (FC) at resting state, requiring n=30 participants per arm (total n=90), allowing for a 10% drop-out rate. Participants were instructed to consume the study product daily, together with breakfast, dissolved in 200 ml of cold or room temperature water or milk. All three study products were provided as indistinguishable maltodextrin-based powders (5 g), packed in identical sachets and provided by AnaBio Technologies. The probiotic products contained 6×10^9^ CFU *Lacticaseibacillus rhamnosus* HN001 with or without food-grade plant-based protein coating (*i.e.* microencapsulation). Consumption compliance was higher than 90% in all groups^20^. Adverse events were fewer in the probiotic groups compared to placebo^20^. Participants were asked to maintain their lifestyle, diet, medications, physical activity and sleep habits throughout the study. The study visits took place at Örebro University, Örebro, Sweden, in two cohorts from March to November 2023. For further details on the intervention study, including questionnaires and missing data handling, please refer to RodeCHutchinson et al. 2025^20^.

### Brainstem-optimised fMRI

Resting state fMRI examinations were conducted with brainstem-optimised protocols for acquisition and analyses. Only full MRI datasets (including data from both before and after intervention of the same participant) were analysed, namely total n=83: placebo (n = 27), encapsulated probiotic (n = 26), and non-encapsulated probiotic (n = 30). A participant flow chart (including reasons for missing data) can be found in Supplementary Figure 1.

Research questions focused on i) within-brainstem FC (ROI-to-ROI connectivity, RRC), and ii) brainstem-to-cortex/subcortex/cerebellum FC (RRC, and seed-based connectivity, SBC). Cortex-to-brainstem FC was not of interest, since the hypothesis was that probiotic effects modulate within-brainstem FC and are transmitted via brainstem to cortex, not *vice versa*.

*A priori* hypothesis-driven selected seeds of interest were: the nucleus tractus solitarius (NTS), as a major integrator of information along the gut-brain afferent pathway^26^; the locus coeruleus (LC), given its connections with the NTS and involvement in the hypothalamic-pituitary-adrenal axis and norepinephrine system^26^; the dorsal raphe nucleus (DRN), as a major serotonergic hub^26^; the ventral tegmental area (VTA), as a key dopaminergic nucleus^26^; and the periaqueductal gray (PAG), as a region shown to be associated with probiotic intervention effects^19^, irritable bowel syndrome^27^, visceral pain^28^ and stress regulation^29^. Other included brainstem regions were: red nucleus (RN), substantia nigra (SN), and subthalamic nucleus (STN). Masks were obtained bilaterally or split into hemispheres as fit.

Baseline resting state brainstem FC was assessed in all n=83 participants and tested for Pearson’s correlation with participant continuous characteristics (age, MoCA, HADS, PSS, PSQI, GSRS, hsCRP); furthermore, FC of male *versus* female participants was contrasted. Additional multiplicity correction by Bonferroni was applied for the number of covariates (eight) tested, resulting in cluster size p-FDR<0.0063 being regarded as significant.

Probiotic effects on resting state brainstem FC were assessed in factorial designs. First, interaction effects, main group and main time effects were assessed. If any of those was significant (p-FDR-corrected<0.05), within-group (before *versus* after intervention) and between-group (before and after intervention) *posthoc* comparisons were evaluated. *Posthoc* comparisons (nine) were additionally Bonferroni-corrected for multiple testing, resulting in cluster size p-FDR<0.0056 being regarded as significant.

For details, see Supplementary Information 2.

## RESULTS

### Study population

Detailed phenotypic participant characterisation, including concomitant medications, is reported in RodeCHutchinson et al. 2025^20^. In brief, sex, age, physical activity and habitual diet at baseline did not differ between groups. Also, cognitive and psychological status (anxiety, depression, stress), sleep quality, gastrointestinal symptoms and inflammatory status did not differ significantly between groups. A shortened summary, specific for the n=83 participants with full datasets, is presented in Table 1.

**Table 1.** Study participant’s baseline characteristics.

|  | **All** | **Placebo** | **Encapsulated probiotic** | **Non-encapsulated probiotic** | **p-value** |
|---|---|---|---|---|---|
| **n** | 83 | 27 | 26 | 30 | - |
| **Sex (f/m)** | 55/28 | 19/8 | 16/10 | 20/10 | 0.7923 |
| **Age<br>(mean±SD, min-max)** | 68.9±5.0, 60-80 | 69.4±5.3, 60-80 | 69.1±5.2, 60-77 | 68.5±4.8, 60-80 | 0.8155 |
| **MoCA score<sup>#</sup><br>(median, min-max)** | 26, 21-30 | 26, 22-30 | 26, 22-29 | 26, 21-28 | 0.8461 |
| **HADS score<sup>#</sup><br>(median, min-max)** | 5, 0-23 | 4, 0-17 | 5, 0-23 | 5, 1-17 | 0.9666 |
| **PSS score<sup>#</sup><br>(median, min-max)** | 10, 1-25 | 10, 1-25 | 11, 3-23 | 9, 2-18 | 0.3345 |
| **PSQI score<sup>#</sup><br>(median, min-max)** | 5, 1-14 | 5, 1-14 | 5, 1-14 | 4, 1-12 | 0.2190 |
| **GSRS score<sup>##</sup><br>(median, min-max)** | 1.5, 1.0-3.1 | 1.5, 1.0-2.5 | 1.5, 1.1-3.1 | 1.5, 1.0-2.6 | 0.8491 |
| **Inflammatory status by hsCRP in plasma [mg/l]<br>(median, min-max)** | 0.98, 0.08-5.29 | 0.96, 0.21-4.95 | 0.96, 0.33-5.29 | 1.43, 0.08-4.81 | 0.4321 |

Abbreviations: MoCA – Montreal Cognitive Assessment, HADS – Hospital Anxiety and Depression Scale, PSS – Perceived Stress Scale, PSQI – Pittsburgh Sleep Quality Index, GSRS – Gastrointestinal Symptom Rating Scale, hsCRP – high sensitivity C-reactive protein. Statistical comparisons between the groups with Chi-square test (sex), one-way ANOVA (age), or Kruskal-Wallis test (MoCA, HADS, PSS, PSQI, GSRS, hsCRP) in GraphPad Prism, version 10.2.2. <sup>#</sup>sum rating of all items, <sup>##</sup>average rating of all items.

### Baseline resting state brainstem functional connectivity

Within-brainstem functional connectivity (FC) at baseline was assessed between all 13 included brainstem structures using ROI-to-ROI connectivity (RRC) (Figure 1A). Overall brainstem-to-cortex/subcortex/cerebellum FC at baseline is presented as a full RRC matrix (Figure 1B) and as region-based connectivity with seeds (Figure 1C). The brainstem FC pattern loaded most strongly and most significantly onto the cerebellum and loaded on all customised networks, except the thalamocortical network (here comprised of bilateral thalamus). For statistical results reporting, see Supplementary Table 3.

**Figure 1.**
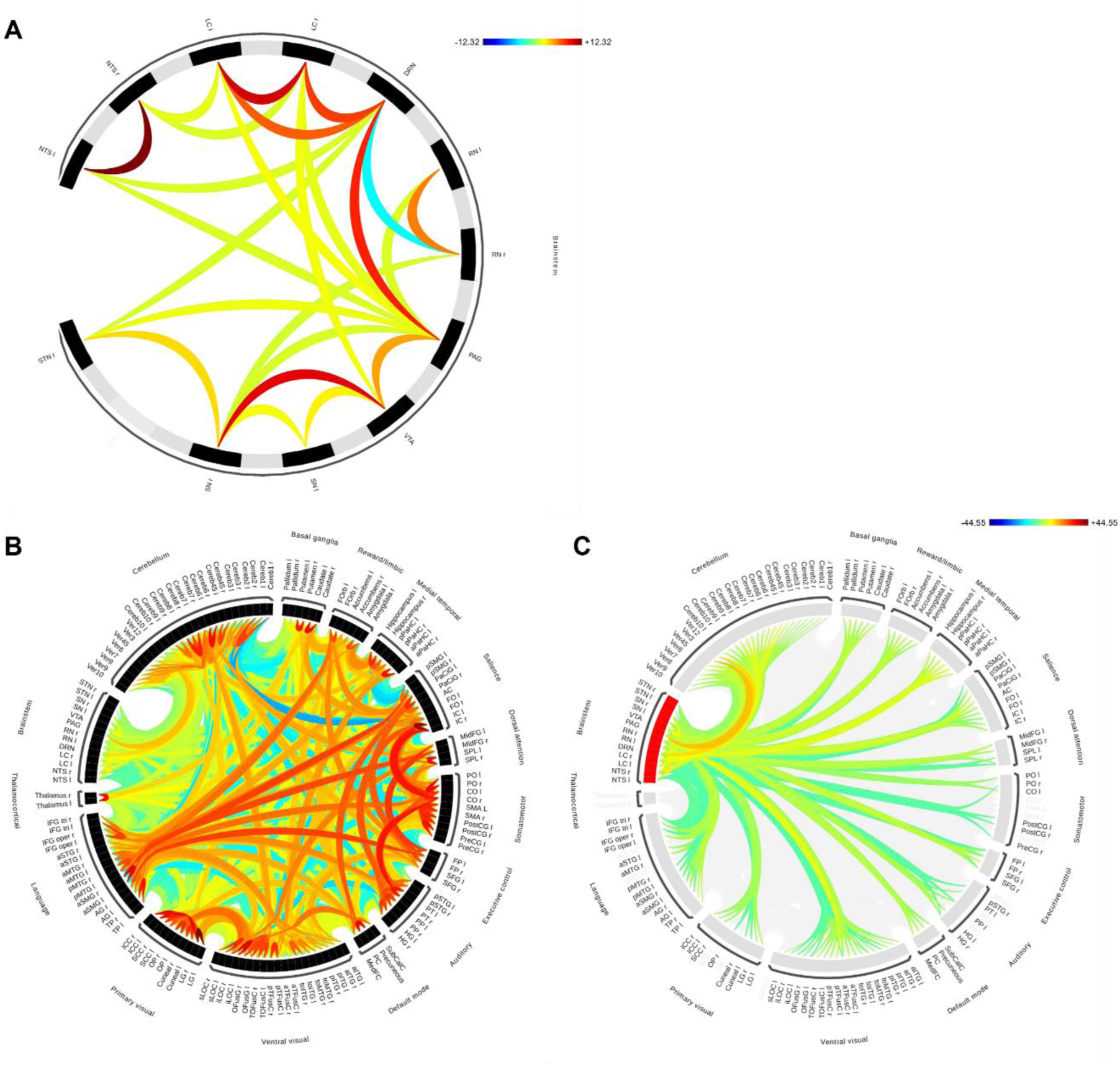
ROI-to-ROI connectivity (RRC) matrix of n=83 healthy elderly at baseline. A within-brainstem functional connectivity (FC), B overall brainstem-to-cortex/subcortex/cerebellum FC, and C region-based brainstem-to-cortex/subcortex/cerebellum FC with brainstem seeds. B and C share the same colour bar. Abbreviations: NTS – nucleus tractus solitarius, LC – locus coeruleus, DRN – dorsal raphe nucleus, RN – red nucleus, PAG – periaqueductal gray, VTA – ventral tegmental area, SN – substantia nigra, STN – subthalamic nucleus, l – left, r – right. For details on groupings of cortical, subcortical and cerebellar regions and explanations of their abbreviations please refer to Supplementary Table 2.

Functional connectivity of the ventral tegmental area (VTA), periaqueductal gray (PAG), dorsal raphe nucleus (DRN), locus coeruleus (LC), and nucleus tractus solitarius (NTS) was furthermore assessed with seed-based connectivity (SBC) (Supplementary Figure 4).

### Factors influencing baseline resting state brainstem functional connectivity

Within-brainstem and overall brainstem-to-cortex/subcortex/cerebellum FC (assessed as RRC) was not affected by any of the assessed covariates (age, sex, Montreal Cognitive Assessment (MoCA), Hospital Anxiety and Depression Scale (HADS), Perceived Stress Scale (PSS), Pittsburgh Sleep Quality Inventory (PSQI), Gastrointestinal Symptom Rating Scale (GSRS), high-sensitivity C-reactive protein (hsCRP)).

Age correlated significantly with the FC of PAG, DRN and LC. Both MoCA and HADS correlated significantly with FC of PAG. PSS correlated significantly with FC of NTS. Furthermore, female and male participants presented significantly different FC of DRN (all SBC with cluster size p-FDR<0.0063 for additional multiplicity correction) (Table 2). None of the assessed baseline factors correlated with FC of VTA. PSQI, GSRS, and hsCRP did not correlate with FC of any of the brainstem seeds.

**Table 2.**
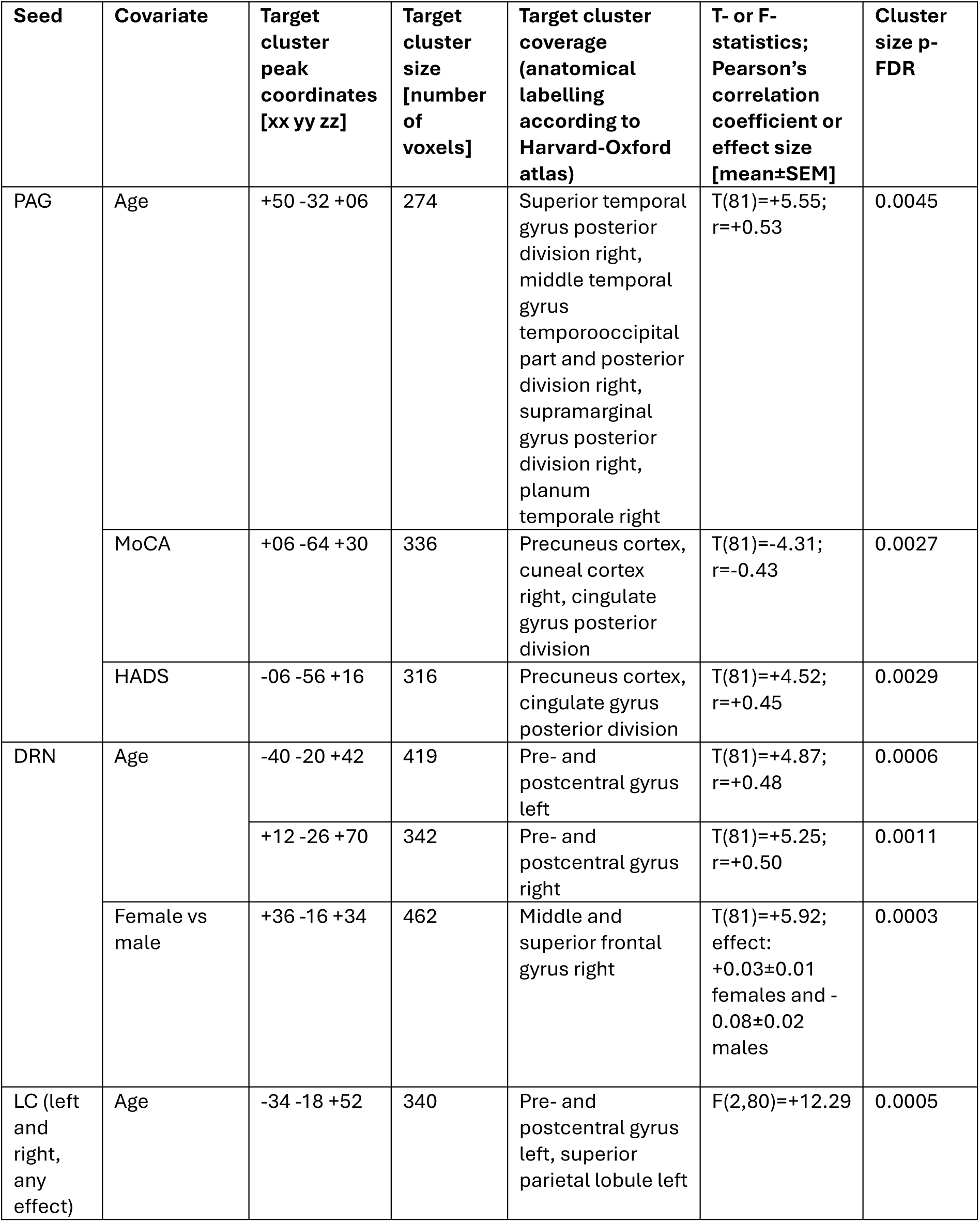
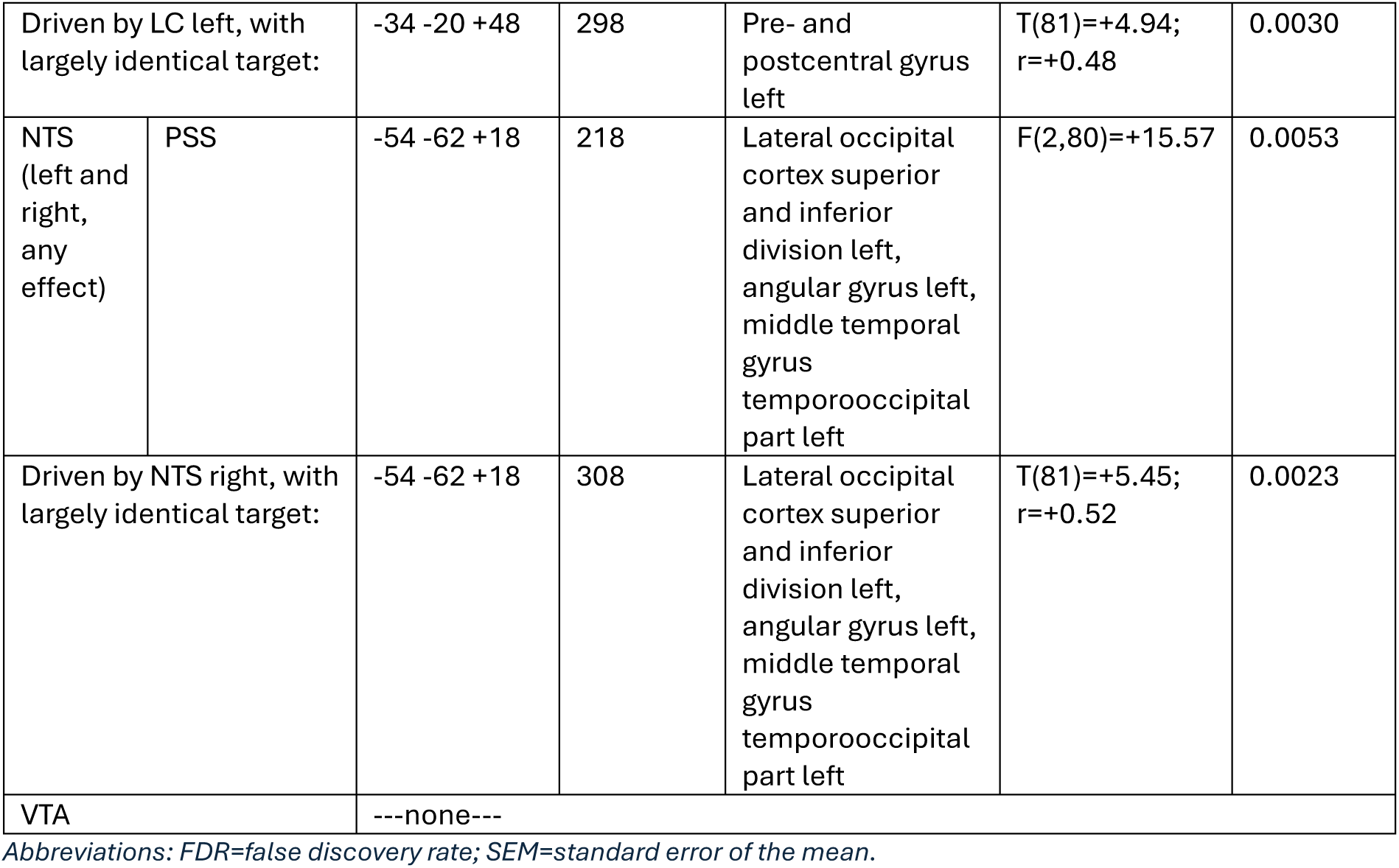
Baseline factors (age, sex, Montreal Cognitive Assessment (MoCA), Hospital Anxiety and Depression Scale (HADS), Perceived Stress Scale (PSS), Pittsburgh Sleep Ǫuality Index (PSǪI), Gastrointestinal Symptom Rating Scale, (GSRS), high sensitivity C-reactive protein (hsCRP)) correlation with functional connectivity of a priori selected brainstem regions of interest (periaqueductal gray (PAG), dorsal raphe nucleus (DRN), locus coeruleus (LC) and nucleus tractus solitarius (NTS), ventral tegmental area (VTA)). Results of seed-based connectivity (SBC); reported if cluster size p-FDR<0.00c3 due to additional multiplicity correction for the number of covariates tested.

| Seed | Covariate | Target cluster peak coordinates [xx yy zz] | Target cluster size [number of voxels] | Target cluster coverage (anatomical labelling according to Harvard-Oxford atlas) | T- or F-statistics; Pearson's correlation coefficient or effect size [mean±SEM] | Cluster size p-FDR |
|---|---|---|---|---|---|---|
| PAG | Age | +50 -32 +06 | 274 | Superior temporal gyrus posterior division right, middle temporal gyrus temporooccipital part and posterior division right, supramarginal gyrus posterior division right, planum temporale right | T(81)=+5.55; r=+0.53 | 0.0045 |
|  | MoCA | +06 -64 +30 | 336 | Precuneus cortex, cuneal cortex right, cingulate gyrus posterior division | T(81)=-4.31; r=-0.43 | 0.0027 |
|  | HADS | -06 -56 +16 | 316 | Precuneus cortex, cingulate gyrus posterior division | T(81)=+4.52; r=+0.45 | 0.0029 |
| DRN | Age | -40 -20 +42 | 419 | Pre- and postcentral gyrus left | T(81)=+4.87; r=+0.48 | 0.0006 |
|  |  | +12 -26 +70 | 342 | Pre- and postcentral gyrus right | T(81)=+5.25; r=+0.50 | 0.0011 |
|  | Female vs male | +36 -16 +34 | 462 | Middle and superior frontal gyrus right | T(81)=+5.92; effect: +0.03±0.01 females and -0.08±0.02 males | 0.0003 |
| LC (left and right, any effect) | Age | -34 -18 +52 | 340 | Pre- and postcentral gyrus left, superior parietal lobule left | F(2,80)=+12.29 | 0.0005 |
| Driven by LC left, with largely identical target: |  | -34 -20 +48 | 298 | Pre- and postcentral gyrus left | T(81)=+4.94; r=+0.48 | 0.0030 |
| NTS (left and right, any effect) | PSS | -54 -62 +18 | 218 | Lateral occipital cortex superior and inferior division left, angular gyrus left, middle temporal gyrus temporooccipital part left | F(2,80)=+15.57 | 0.0053 |
| Driven by NTS right, with largely identical target: |  | -54 -62 +18 | 308 | Lateral occipital cortex superior and inferior division left, angular gyrus left, middle temporal gyrus temporooccipital part left | T(81)=+5.45; r=+0.52 | 0.0023 |
| VTA |  | ---none--- |  |  |  |  |

*Abbreviations: FDR=false discovery rate; SEM=standard error of the mean.*

### Probiotic intervention effects on resting state brainstem functional connectivity

Within-brainstem and brainstem-to-cortex/subcortex/cerebellum RRC did not show any significant intervention effects (*i.e.* no main group, main time, or time*group interaction effects).

Assessing FC of the VTA (with SBC), a significant time*group interaction effect was observed with a target cluster covering precuneus, right intra-and supracalcarine cortex and lingual gyrus (Table 3). *Posthoc* analysis revealed that there was a significant within-group change from before to after intervention in the non-encapsulated probiotic group with a target cluster covering left occipital fusiform gyrus, temporal occipital fusiform cortex, lateral occipital cortex inferior division and cerebellum 6 and crus1. No significant within-group effects were observed in the placebo or encapsulated probiotic groups, and no significant between-group differences were observed before or after intervention.

**Table 3.** Functional connectivity of one a priori selected brainstem region of interest, namely ventral tegmental area (VTA). Results of seed-based connectivity (SBC); 2×3 factorial design results (i.e. main group, main time, or time*group interaction effects) reported if cluster size p-FDR<0.05, posthoc test results reported if cluster size p-FDR<0.005c due to additional multiplicity correction for the number of multiple comparisons tested.

| **Seed** | **Type of effect** | **Target cluster peak coordinates [xx yy zz]** | **Target cluster size [number of voxels]** | **Target cluster coverage (anatomical labelling according to Harvard-Oxford atlas)** | **T- or F-statistics; effect size [mean±SEM]** | **Cluster size p-FDR** |
|---|---|---|---|---|---|---|
| VTA | Time*group interaction effect | +06 -60 +12 | 201 | Precuneus, intra- and supracalcarine cortex right, lingual gyrus right | F(2,80)=-13.15 | 0.0130 |
|  | Non-encapsulated probiotic, intervention effect | -28 -76 -20 | 209 | Occipital fusiform gyrus left, temporal occipital fusiform cortex left, lateral occipital cortex inferior division left, cerebellum 6 and crus1 | T(29)=+5.78; -0.07±0.02 before and +0.04±0.02 after intervention | 0.0055 |

*Abbreviations: FDR=false discovery rate; SEM=standard error of the mean.*

When assessing FC of the LC (left and right, with SBC) a significant main time effect was observed (Table 4). *Posthoc* analyses revealed that there were significant within-group changes from before to after intervention in the non-encapsulated probiotic group. Those within-group changes were dominated by FC of the right LC with a largely identical target cluster covering right insular cortex, central opercular cortex, putamen, precentral gyrus and planum polare and frontal operculum cortex and another largely identical target cluster in the left insular cortex and Heschl’s gyrus, although not significant after multiplicity correction. Also, within-group changes of only the right LC were observed with a target cluster covering left pre-and postcentral gyrus and superior parietal lobule, largely overlapping with the main time effect, as well as with a target cluster of the left and right LC (though the latter did not remain significant after additional multiplicity correction). Also, the right LC’s target cluster covering left occipital pole and occipital fusiform gyrus, lingual gyrus, and cerebellum crus1 and crus2 overlapped largely with a target cluster of the left and right LC that did not remain significant after additional multiplicity correction. Functional connectivity of the left LC was not affected by the intervention. No significant within-group effects were observed in the placebo or encapsulated probiotic groups, and no significant between-group differences were observed before or after intervention.

**Table 4.**
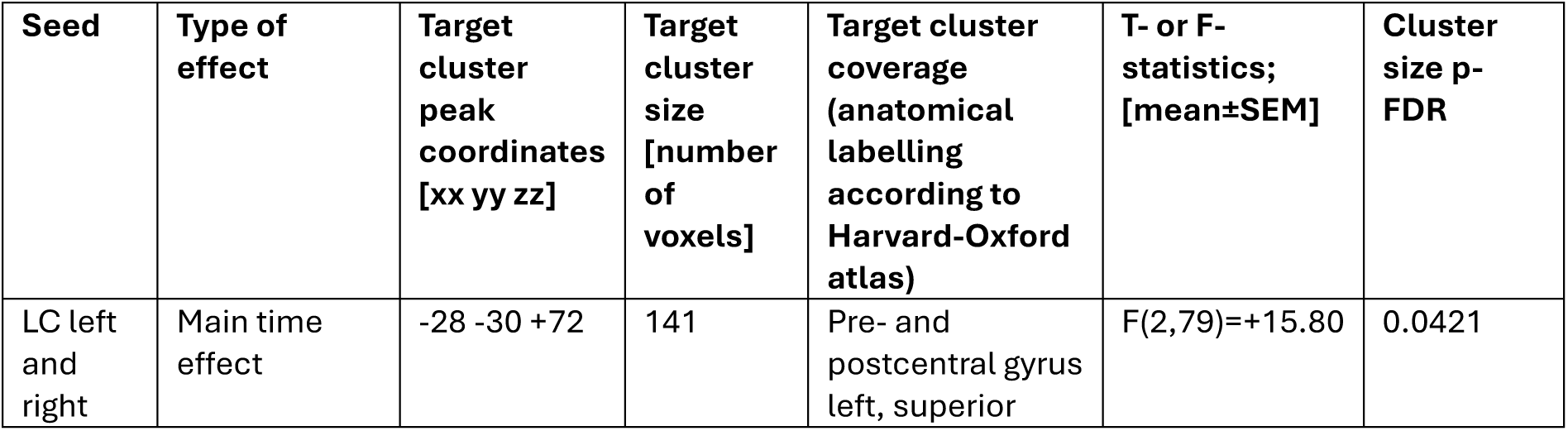
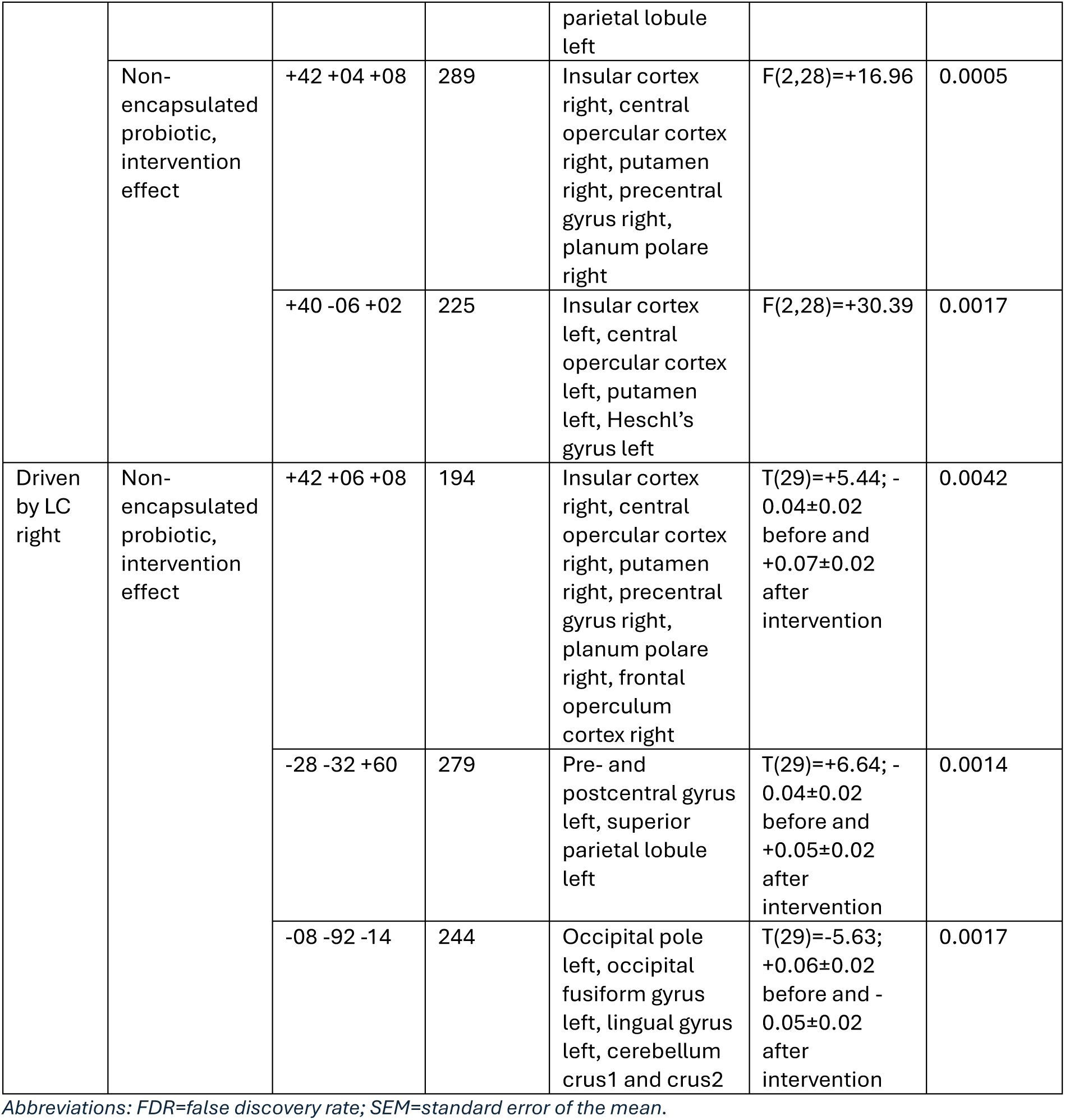
Functional connectivity of one a priori selected brainstem region of interest, namely locus coeruleus (LC). Results of seed-based connectivity (SBC); 2×3 factorial design results (i.e. main group, main time, or time*group interaction effects) reported if cluster size p-FDR<0.05, posthoc test results reported if cluster size p-FDR<0.005c due to additional multiplicity correction for the number of multiple comparisons tested.

Functional connectivity of DRN, PAG, and NTS was not affected by the probiotic intervention (no significant main time, main group, or time*group interaction effects in SBC analyses; hence, no *posthoc* tests were performed).

## DISCUSSION

This work is the first to assess probiotic intervention effects with brainstem-optimised functional magnetic resonance imaging (fMRI) acquisition and analysis. Furthermore, it is one of the first to assess functional connectivity (FC) of the brainstem in a resting state condition in an elderly cohort and, to the best of our knowledge, with the largest sample size thus far. Given the anatomical and functional complexity and heterogeneity of the brainstem, the investigation focused on distinct brainstem nuclei/regions. This study investigated FC patterns among various brainstem regions as well as between brainstem regions and cortical, subcortical and cerebellar regions.

At baseline, FC of the periaqueductal gray (PAG), dorsal raphe nucleus (DRN), locus coeruleus (LC) and nucleus tractus solitarius (NTS) to cortical, but not cerebellar, regions correlated with several phenotypic characteristics. As such, age correlated positively with FC of LC (predominantly left LC) and DRN with pre-and postcentral gyrus (*i.e.* sensorimotor regions), as well as of PAG with temporal gyrus, supramarginal gyrus and planum temporale. These findings are consistent with previous evidence from adults up to the age of 49 years demonstrating age-related alterations in LC connectivity, including increased FC with frontoparietal and cerebellar regions, as well as decreased FC with visual regions^3^. Similarly, ageing has also been shown to play a potential role in weakening brainstem-cortical synchronisation, which was also associated with poorer working memory performance^6^. In addition to correlations with age, in the present study, FC of PAG with precuneus cortex and cingulate gyrus correlated negatively with higher cognitive status (high Montreal Cognitive Assessment (MoCA) score) and better mood (low Hospital and Anxiety Scale (HADS) score), suggesting that brainstem-cortical FC may also vary due to individual differences in cognitive performance and psychological state (Figure 2). Of note, altered FC between midbrain/brainstem nuclei and cortical regions involved in affective processing have been reported in major depressive disorder, including reduced VTA-dorsal anterior cingulate cortex FC, which was associated with greater depressive symptom burden^30^. Moreover, here, perceived stress correlated positively with the FC between NTS (predominantly right NTS) and occipital cortex, angular gyrus and temporal gyrus. Functional connectivity of the DRN differed between men and women. Sleep quality, gastrointestinal symptoms and inflammatory status seemed not to influence brainstem FC. Although the baseline associations of the present study suggest that FC of the brainstem and cortical networks may vary with several phenotypic characteristics implicated in healthy ageing, these cross-sectional associations do not establish conclusions on causality, and hence potential relationships should be further explored in future studies.

**Figure 2.**
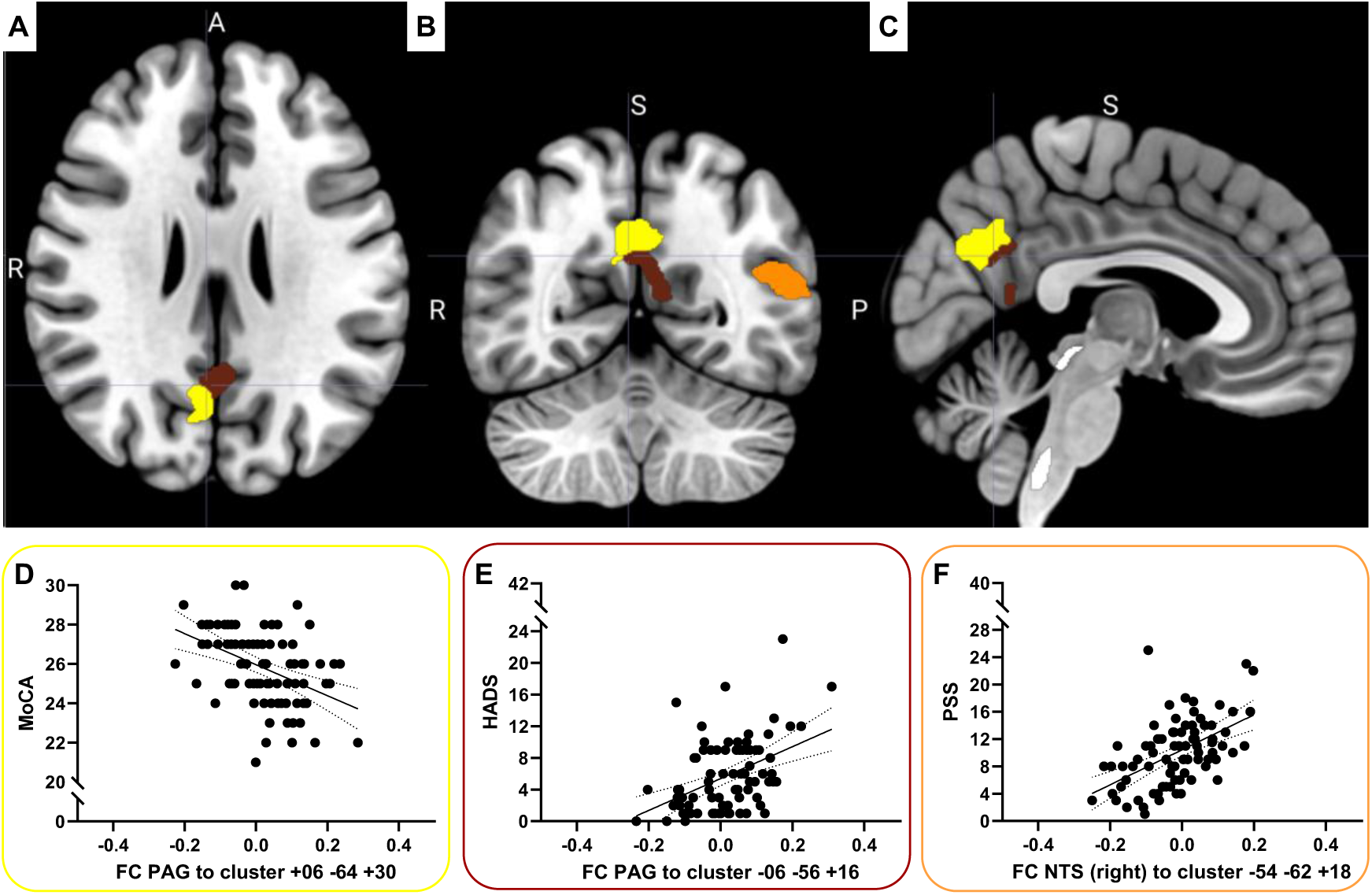
Brainstem-to-cortex functional connectivity (FC) at rest correlated with cognitive performance and psychological symptoms in healthy elderly. A) Axial, B) coronal, and C) sagittal view of brain regions superimposed on a standard brain template in Montreal Neurological Institute (MNI) space. White regions show brainstem seeds. Functional connectivity of the periaqueductal gray (PAG, dorsal white) with the precuneus cortex, right cuneal cortex, cingulate gyrus posterior division (yellow) and the precuneus cortex, cingulate gyrus posterior division (brown) correlated with D) cognitive performance assessed by the Montreal Cognitive Assessment (MoCA) and E) mood assessed by the Hospital Anxiety and Depression Scale (HADS). Functional connectivity of the right nucleus tractus solitarius (NTS, ventral white) with the left lateral occipital cortex superior and inferior division, angular gyrus, middle temporal gyrus temporooccipital part (orange) correlated with stress assessed by the Perceived Stress Scale (PSS). For MoCA the higher the score, the better the cognitive performance, for HADS and PSS the lower the score the better the mood and lower the stress perception. The solid line in the graphs represents the best-fit line of a simple linear regression, the doted lines indicate its S5% confidence bands. n=83 participants. Seed-to-voxel analyses (SBC) performed in CONN Toolbox version 25.b. Figure created in MRIcroGL and GraphPad Prism.

While the brainstem, and more specifically brainstem nuclei such as the NTS, are known as integrators of information along the gut-brain sensory pathway^26^, few studies have assessed this pathway in humans. In the presented study, surprisingly, probiotic intervention did not affect FC of NTS, DRN or PAG. Instead, resting state FC of LC and ventral tegmental area (VTA), were affected. Interestingly, none of the within-group non-encapsulated probiotic intervention effects were simple changes in magnitude of FC but mostly changes from anti-correlated to correlated activation patterns. Both LC and VTA showed altered FC with a cluster covering occipital pole and cortex, amongst others. This cluster includes regions that are involved in high order visual and integrative processing and has also previously shown altered FC with the inferior frontal gyrus and supramarginal gyrus, notably following the same probiotic intervention^20^. Hence, the herein presented findings extend those previously reported by identifying a brainstem-related component possibly contributing to the integration of probiotic-gut-brain signalling. Furthermore, FC of LC with left pre-and postcentral gyrus and superior parietal lobule – a pattern that was also correlated with age (Figure 3) – was affected, together suggesting that the observed intervention effects may be relevant to understanding brainstem-cortical changes during healthy ageing. Also, FC of LC with a cluster covering insular and opercular cortex, putamen and planum polare (*i.e.* regions involved in interoception, emotions, and language processing) was altered following probiotic intervention. This may suggest that probiotic effects and the integration of related gut-derived signals to upstream cortical processes are specific rather than widespread.

**Figure 3.**
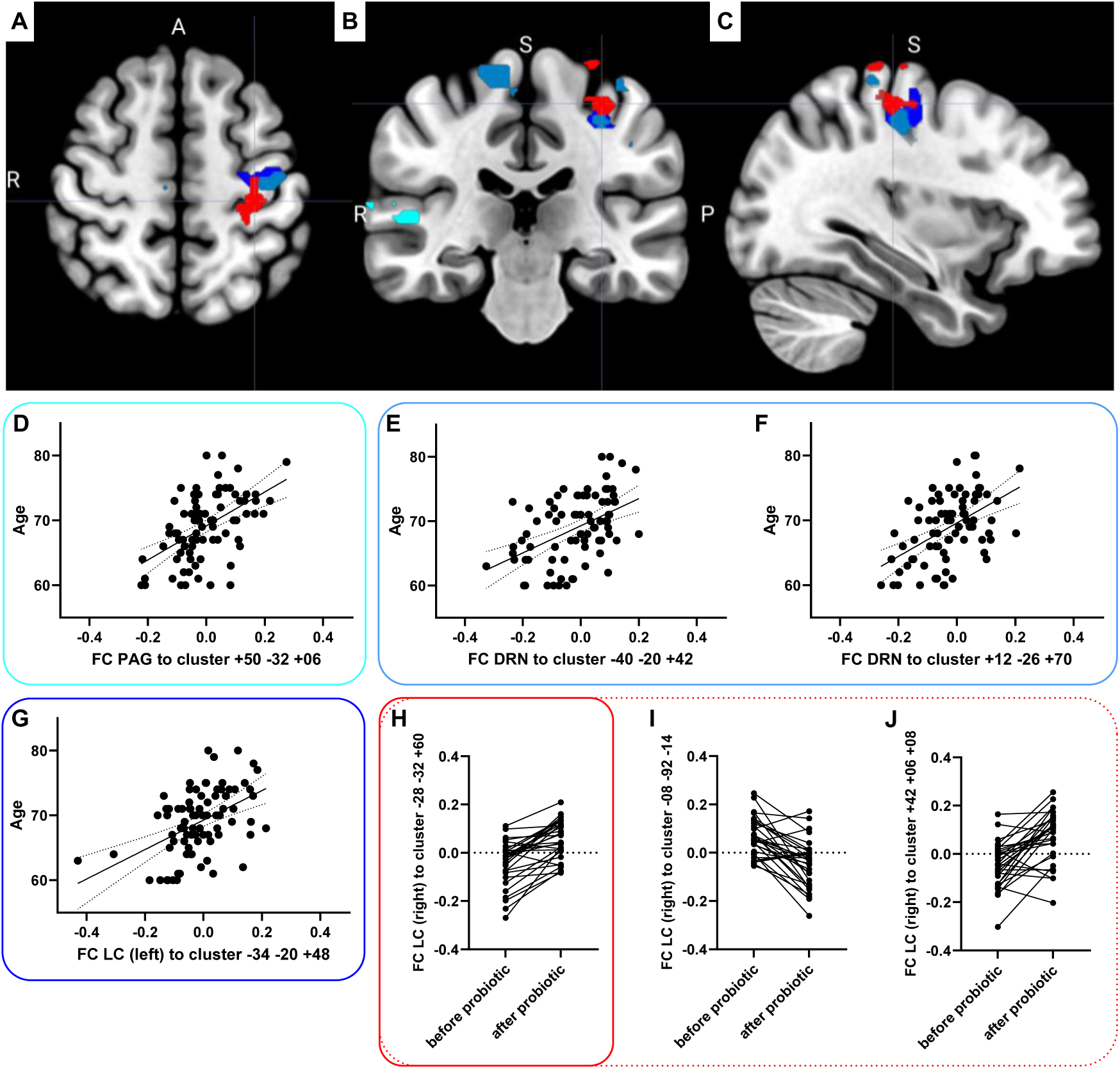
Brainstem-to-cortex functional connectivity (FC) at rest correlated with age and was affected by probiotic intervention healthy elderly. A) Axial, B) coronal, and C) sagittal view of brain regions superimposed on a standard brain template in Montreal Neurological Institute (MNI) space. Functional connectivity of D) the periaqueductal gray (PAG) with the right superior temporal gyrus posterior division, middle temporal gyrus temporooccipital part and posterior division, supramarginal gyrus posterior division, planum temporale (light blue), E and F) the dorsal raphe nucleus (DRN) and bilateral pre-, postcentral gyrus (middle blue), and G) left locus coeruleus (LC) with left pre-, postcentral gyrus (dark blue) correlated with age. H) A largely overlapping cluster covering left pre-, postcentral gyrus, superior parietal lobule (red) showed FC with right LC impacted by non-encapsulated probiotic intervention (illustration of the posthoc within-group effect, for visualisation of the main time effect please refer to Supplementary Figure 5). I and J) show intervention-related changes in FC of the right LC with two other clusters, namely one covering left occipital pole, occipital fusiform gyrus, lingual gyrus, cerebellum crus1 and 2, and the other covering right insular cortex, central opercular cortex, putamen, precentral gyrus, planum polare, frontal operculum cortex (both are not visual in this view). The solid line in the graphs represents the best-fit line of a simple linear regression, the doted lines indicate its S5% confidence bands. n=83 participants totally, n=27 participants received the placebo, n=2c the encapsulated and n=30 the non-encapsulated probiotic intervention. Seed-to-voxel analyses (SBC) performed in CONN Toolbox version 25.b. Brainstem seeds are not visual in this view (see Supplementary Figure 2 and 3). Figure created in MRIcroGL and GraphPad Prism.

To date, there are no published studies explicitly assessing the functional integration of probiotic effects via the brainstem^16^. The first probiotics fMRI study investigating a four-weeks intervention with a fermented milk multi-strain product containing *Bifidobacterium animalis* subsp *lactis*, *Streptococcus thermophilus*, *Lactobacillus bulgaricus*, and *Lactococcus lactis* subsp *lactis* observed altered FC in a network including the PAG^19^. However, it is unclear whether the entire brainstem or solely midbrain/upper brainstem was acquired, and the study seems not to have employed brainstem-specific acquisition and processing techniques^19^. Also, the inclusion of one large brainstem region as per the Harvard-Oxford atlas in our previous whole-brain analyses did not reveal any significant probiotic intervention effects^20^. However, this atlas brainstem region is large and characterised by functional heterogeneity and does not cover the entire brainstem, which may have diluted the findings – hence those findings do not contradict the herein reported ones. Apart from human studies, interestingly, a four-weeks probiotic intervention with *Lactobacillus acidophilus* and *Bifidobacterium bifidum* reduced iron deposition in the substantia nigra of a Parkinson’s disease rat model^31^.

Several animal studies have investigated how anatomical location and type of intestinal stimuli influence which brainstem regions are involved in gut-brain communication. In the small intestine, specialised enteroendocrine cells, referred to as neuropod cells, form synaptic connections with vagal sensory neurons and can rapidly transmit information from the intestinal lumen to the brain; these vagal sensory neurons in turn project to the NTS^32^. Also, sensory and motor pathways from the small intestine substantially overlap in neurons of multiple brain regions, including the NTS and the dorsal vagal complex^33^. This evidence suggests that nutrient sensing in the small intestine involves communication along fast neural synapses rather than solely relying on slower signalling via gut hormones^32^. Also, vagal afferents are densest in the small intestine and contribute strongly to nutrient sensing and sensory signalling^34^ ^35^. Furthermore, signals from the small intestine seem to be integrated via a widespread network rather than a single neural pathway. In the distal colon and rectum, splanchnic and pelvic afferents communicate gut-derived information via distinct pathways and brainstem regions^36^. Surgical removal of splanchnic afferent neurons reduced responses to colorectal distension in the caudal and rostral ventromedial medulla and the lateral parabrachial nucleus, whereas removal of pelvic afferent neurons reduced responses in the rostral ventromedial medulla, lateral parabrachial nucleus, LC, Barrington’s nucleus, and PAG^36^. Taken together, these findings indicate that the brainstem regions involved in processing gut-derived information may depend on both the location of the intestinal signal and the type of sensory information (nutrient sensing, noxious stimuli, etc.) being transmitted. This is especially important since the two probiotic product formulations may act via different mechanisms, with the micro-encapsulation intended (but not measured in this study) to support gastrointestinal survival and reach to more distal parts of the gastrointestinal tract. While no between-group differences were observed, the within-group changes may indicate that the non-encapsulated probiotic may act via neuronal gut-brain signalling and the encapsulated probiotic via other pathways.

In addition, human studies have also examined the pathways by which location and type of intestinal stimuli differentially engage brainstem regions. For example, duodenal infusion of capsaicin, an intestinal chemonociceptive stimulus, resulted in increased activation in the NTS compared to placebo; this activation was independent of reported pain^11^. In another study, ingestion of high-*versus* low-fat yogurt altered NTS connectivity with regions involved in reward and gustatory networks, including insula, temporal cortex, caudate, putamen, frontal operculum, and globus pallidus^12^. Also, a high-sucrose beverage increased power in the highest frequency band (slow-3) in the NTS and reduced low frequency power (slow-5) in several brainstem regions, including the LC, PAG, and pontine/trigeminal nuclei compared to a low-sucrose beverage with identical sweetness^13^, providing evidence that nutrient ingestion can affect both the NTS as well as other brainstem regions. This distinction is relevant to the herein presented study in which *Lacticaseibacillus rhamnosus* HN001 was associated with changes in FC involving the LC and VTA. The NTS acts as a brainstem relay for visceral sensory information, particularly signals via vagal afferents. However, the absence of intervention-related changes in NTS connectivity does not necessarily preclude effects on other brain circuits. The changes in LC connectivity following *Lacticaseibacillus rhamnosus* HN001 supplementation may instead reflect modulation of brainstem pathways beyond the NTS. However, the present findings cannot determine whether the LC was influenced through the NTS, through other visceral sensory pathways, or through indirect gut-brain signalling mechanisms.

Importantly, interpretation of findings should be within the context of methodological complexity. We employed a brainstem-optimised fMRI acquisition with complete coverage of brain and brainstem, hence a larger field of view, enabled by integrated utilisation of head and bed coils. Simultaneously, high temporal resolution was maintained, using simultaneous multi-slice sampling. Our usual approach for motion minimisation, involving mild immobilisation with foam pads performed by experienced nurses, has previously been shown effective^20^.

Subsequently, we employed brainstem-optimised analyses. Effects of physiological noise (e.g. cardiac and respiratory), including effects of their aliased signals, were reduced i) by using bandpass filtering, and ii) based on signals from the eroded white and cerebrospinal fluid (CSF) maps, especially important due to their anatomical proximity to brainstem regions, by denoising including aCompCor. Cardiac and respiratory recordings have not been collected, and while its inclusion as covariates could have improved signal-to-noise ratio (SNR), its exclusion can be regarded as more conservative by underestimating rather than overestimating FC^37^. Nonetheless, future studies should consider collecting simultaneous physiological recordings (e.g. heart and breathing rate) in order to assess organ response to autonomic signalling. Also, scrubbing has been applied, including removal of nearby volumes. Brainstem regions have been masked for grey matter and have been analysed at their acquired voxel size. Resampling into smaller voxel sizes has been deemed obsolete since it simply would have divided the signal into a smaller grid. Smoothing has not been applied for brainstem regions in order to preserve their small volumes, although at the likely cost of a lower SNR. The resulting 2.5 mm^3^ voxel size is comparable with other studies^6^ ^9^ ^11^ ^30^. Some newer studies with smaller voxel sizes are performed at 7T, which however comes with its own challenges; moreover, findings are often confirmed at 3T^38–40^ (as also used in the presented setting).

Standard FDR-based multiplicity correction approaches have been applied per analysis. Furthermore, the cross-sectional analyses have been additionally multiplicity-corrected for the evaluation of multiple correlations, and the intervention effect *posthoc* analyses were additionally multiplicity-corrected for the number of comparisons as standard for ANOVAs, both using Bonferroni correction. Exact p-values were reported, allowing the reader to apply further correction if desired. This approach aligns well with the exploratory character of the presented work.

The herein investigated elderly cohort presented a variation in cognitive status, sleep quality, and inflammatory status, all with their medians clustering around the cut-off values that indicate mild cognitive impairment (MoCA<26), impaired sleep (PSQI>5), or increased inflammation (hsCRP>1 mg/l). The cohort also presented variation in mood and perceived stress, with the scores reaching beyond the cut-offs (HADS>11, PSS>13), but the median remained within the normal range. Participants presented variation on the GSRS with none to mild gastrointestinal symptoms (GSRS<4). Also, age spanned across the entire inclusion range of 60–80 years of age, with a median of69 years of age. All in all, this supports the external validity and generalisability of the results, although confirmation in a larger sample is warranted. Furthermore, the included baseline characteristics were suitable for correlation analyses which require some degree of variation. Importantly, none of the characteristics differed between groups at baseline, confirming that the randomisation to the intervention arms was successful. Therefore, none of the baseline factors was corrected for during intervention effect analyses. Given the strong age effect, age distribution and stratification should be kept in mind also in future studies. The herein presented findings further provide rational to investigate individuals with mental health issues, cognitive impairment or disorders of gut-brain interactions.

The presented study employed a probiotic intervention as a physiological model. From a gut-brain perspective, imaging the brainstem adds to the understanding of (patho-)physiological processes and modes-of-action of intervention effects, and may hence have clinical implications especially in the perspective of neurodegenerative disorders. Functional connectivity of the brainstem and cortical networks may vary with individual phenotypic characteristics implicated in healthy ageing and may be further modified by probiotics.

## ETHICS STATEMENT

The study was performed according to the Declaration of Helsinki and its revisions, approved by the Swedish Ethical Review Authorities (original approval on date 2023-01-30, registration number 2022-06042-01, and amendment 2025-02584-02) and registered at ClinicalTrials.gov under ID: NCT05801042 (submission date 2023-02-10). All participants received written and verbal information and provided written informed consent before any study-related activity. Participation was voluntary and could be withdrawn at any time for any reason without needing to disclose the reason. Study participants received 2000 SEK (taxable income), compensating for time and potential discomfort.

## DATA AVAILABILITY STATEMENT

The data are pseudonymised, and the key variable may not be destroyed. According to Swedish ethics regulations, the raw data cannot be shared without an approved ethics application from the National Swedish Ethics Authority. An ethical permit can only be obtained for research being conducted within Sweden. Data can be requested as a public document via and will have to undergo a confidentiality assessment to determine what can be released.

## PUBLIC AND PATIENT INVOLVEMENT

Participants have not been involved in design and study execution, yet apprehensible results of the main study have been disseminated to them. Part of the group-level results have been presented to healthcare professionals and scientific audiences.

## ACKNOWLEDGEMENTS

We thank the staff at the Center for Experimental and Biomedical Imaging in Örebro (CEBIO) for the conductance of MRI investigations, Myrto Chatzopoulou and other former and current colleagues at the Nutrition-Gut-Brain Interactions Research Centre for assistance during data collection. The authors acknowledge AnaBio Technologies for development, manufacture and provision of the probiotic and placebo study products and for technical support relating to the study formulations.

## FUNDING

This study was supported by AnaBio Technologies, Dublin, Ireland. Study team and sponsor have been in regular exchange, yet, the sponsor did not have any influence on the analyses nor interpretation. This study was furthermore conducted within the Responsive Nutrition Profile Rosetta@ORU, funded by the Swedish Knowledge Foundation (grant number KKS 20230004-H-01).

## COMPETING INTERESTS

None of the authors have any conflicts of interest to declare.

## CRediT AUTHORSHIP CONTRIBUTION STATEMENT

Julia Rode: Writing – review C editing, Writing – original draft, Visualisation, Validation, Supervision, Project administration, Methodology, Investigation, Formal analysis, Data curation, Software, Conceptualisation.

Ashley N Hutchinson: Writing – review C editing, Writing – original draft, Visualisation, Validation, Supervision, Project administration, Investigation, Conceptualisation.

Per Thunberg: Writing – review C editing, Writing – original draft, Validation, Methodology, Investigation, Software, Resources, Conceptualisation.

Robert J Brummer: Writing – review C editing, Supervision, Resources, Investigation, Funding acquisition, Conceptualisation.

## Notes

### Clinical Trial

NCT05801042

### Author Declarations

The study was approved by the Swedish Ethical Review Authorities (original approval on date 2023-01-30, registration number 2022-06042-01, and amendment 2025-02584-02).

