## Supplementary Information for "Brainstem functional connectivity integrates microbiome-gut-brain signals in healthy older adults: a cross-sectional evaluation and a randomised controlled probiotic trial"

*Supplementary Information 1.*

High-sensitivity C-reactive protein (hsCRP) analysis

Morning blood samples (Li-heparin plasma) were collected after overnight fasting, stored at -80°C until analysis using the Cardiophase hsCRP assay (REF 06837459, Siemens Healthcare, Germany) and the Siemens ADIVA 1800 Chemistry System (Siemens Healthcare) according to hospital routines at the laboratory of clinical chemistry, Örebro University Hospital, Örebro, Sweden.

*Supplementary Information 2.*

Brainstem-optimised functional magnetic resonance imaging (fMRI) acquisition and analyses

All MRI examinations were conducted at similar times between 8 am and 1 pm at the Center for Experimental and Biomedical Imaging in Örebro (CEBIO) to reduce potential variation related to time of day. Participants were asked to follow the same routines on both days of MRI examinations, including avoidance of physical exercise and limiting the consumption of coffee or tea to one cup if they'd usually drink some in the morning.

A 3.0 T MR system (Signa Premier, GE Medical Systems, WI) was utilised with both a 48-channel head coil and spine coils integrated within the table. An initial structural scan (3D T1w IR-prepared fast spoiled gradient recalled echo, "BRAVO") of 3 min and 50 seconds min was performed, followed by a fluid attenuated inversion recovery (3D cube FLAIR) scan of 4 min and 34 seconds, and an eyes-open resting state fMRI scan (2D gradient echo EPI pulse sequence) of 10 min. A mirror was mounted on the head coil, allowing the study participants to view a monitor during image acquisition. There, during the resting state scan, a white fixation cross was displayed on a black background.

A fully trained and experienced MRI nurse and a qualified investigator conducted the MRI session and were present at all times. Foam pads provided comfort and mild immobilisation. Lights in the room were switched off during resting state fMRI.

Parameters for the T1-weighted structural scan were: TR/TE/TI = 7.3/3.0/450 ms, flip angle = 12°, acquired voxel size = 0.9×0.9×1.2 mm, parallel imaging acceleration (ARC) factor of 2, Field-of-View (FoV) = 240 mm, number of axial slices = 316 and an image matrix of 256×256.

The following parameters were applied for the FLAIR acquisition: TR/TE/TE = 6300/104/1849 ms, parallel imaging (ARC) = 2×2, hypersense = 1.2, echo train length = 180, acquired voxel size = 1×1×1.2 mm, 320 sagittal slices and phase encoding in the anterior-posterior (AP) direction.

Resting state fMRI was acquired with axial interleaved slice order, number of slices = 76, slice thickness = 2.5 mm, pixel size = 2.5×2.5 mm, no slice gap, FoV = 240 mm, flip angel = 90°, TR/TE = 2500/35 ms, fat saturation, phase-encoding in AP-direction, ARC factor of 2 and a hyperband factor of 2. One resting state fMRI scan contained 240 volumes. In addition, four prior dummy scans were also acquired to obtain steady-state tissue magnetisation; however, these were not included in the analyses.

The FoV and number of slices were sufficient to ensure complete coverage of the brain and of the brainstem down to at least the second cervical vertebra (C2), in some cases down to C3. Both the head coil and the table coils were connected through AutoSelect, implying that if the participant's brainstem due to anatomical reasons ended up outside the head coil, coil elements from the spine coil were automatically activated to ensure that both brain and brainstem were encompassed.

Acquired images were converted to the nifti-format using dcm2niix (https://github.com/rordenlab/dcm2niix), and data was subsequently preprocessed as described in Rode&Hutchinson et al. 2025[1] using CONN Toolbox[2]. In brief, it included realignment, slice-time correction, segmentation and normalisation into Montreal Neurological Institute (MNI) space, ART-based outlier detection for scrubbing including removal of nearby volumes (motion outlier detection at a framewise displacement threshold of 0.9 mm and/or a global BOLD signal z-score threshold of 5 at timepoint t), and denoising including linear detrending, the aCompCor routine (five white matter and cerebrospinal fluid (CSF) components) and low frequency band-pass filtering at 0.008 to 0.09 Hz, accounting, amongst others, for physiological noise. Data was quality controlled, and, on average, <1.1% of volumes per participant and session were removed. All processing and analyses specific to the herein presented work were performed in CONN Toolbox (version 25.b, using Matlab R2025a).

Binary brainstem masks (Supplementary Table 1) of subthalamic nucleus (STN), substantia nigra (SN), ventral tegmental area (VTA), periaqueductal gray (PAG), red nucleus (RN), dorsal raphe nucleus (DRN), locus coeruleus (LC) and nucleus tractus solitarius (NTS) were obtained from the probabilistic Levinson Bari atlas[3] applying thresholds for 10–90% (downloaded from https://www.uclahealth.org/departments/neurosurgery/research/research-grant-funding/surgical-neuromodulation-and-brain-mapping-lab/research-areas) and PD25 atlas[4] (downloaded from https://nist.mni.mcgill.ca/multi-contrast-pd25-atlas/). Masks were obtained bilaterally (VTA, PAG, DRN) or split into left and right hemispheres (STN, SN, RN, LC, NTS) and integrated into the analyses applying individual participant grey matter masking to account for false signals from nearby structures, such as white matter or CSF.

Successful registration of those brainstem masks was determined on a random selection of several individual participant's normalised structural images. Due to data integrity reasons such individual registrations are not shown; instead, the registration is visualised on mean structural images (Supplementary Figures 2 and 3).

ROI-to-ROI connectivity (RCC) and seed-based connectivity (SBC) analyses were performed. To account for the small structures of the brainstem, RRC was based on unsmoothed data. The BOLD signal of seeds in the SBC was extracted from unsmoothed data and correlated with smoothed data on voxel level. Spatial smoothing was applied using a Gaussian kernel of 8x8x8 mm; this hence applies to cortical, subcortical and cerebellar regions.

Within-brainstem RRC was conducted with all 13 imported brainstem masks (VTA, DRN, PAG, RN L/R, SN L/R, STN L/R, LC L/R, NTS L/R). Brainstem-to-cortex/subcortex/cerebellum RRC was conducted for those and all 131 Harvard-Oxford atlas regions (all those available in CONN except the large brainstem ROI). The latter RRC was also visualised as region-based connectivity with seeds, not changing the statistical approach from default RRC but the visualisation. For groupings and explanations of labels see Supplementary Table 2. SBC used a subset of the imported brainstem masks as seeds and targeted any voxel. Seeds of interest were: NTS, LC, DRN, VTA and PAG.

First-level (within subject) and second-level (between subjects) analyses were performed with the following settings on first-level: bivariate correlation, HRF weighting, source dimensions: 1, no temporal expansion, no frequency decomposition; and on second-level: cluster-based inference #1 functional network connectivity with cluster threshold $p<0.05$ cluster-level p-FDR-corrected (multivariate omnibus test), connection threshold $p<0.05$ p-uncorrected for RRC; and cluster-based inference #1 random field theory parametric statistics with cluster threshold $p<0.05$ cluster-size p-FDR-corrected, voxel threshold $p<0.001$ p-uncorrected for SBC.

Baseline (*i.e.* before intervention) resting state brainstem functional connectivity was assessed in all n=83 participants (between-subjects contrast [1]) and tested for correlation (between-subjects contrast [0 1]) with participant continuous characteristics (age, MoCA, HADS, PSS, PSQI, GSRS, hsCRP), connectivity of male versus female participants was contrasted [-1 1]. Additional multiplicity correction by Bonferroni was applied for the number of covariates (eight) tested, resulting in cluster size p-FDR<0.0063 being regarded as significant.

Probiotic effects on resting state brainstem functional connectivity were assessed in factorial designs with three groups (*i.e.* encapsulated, non-encapsulated probiotic, placebo) and two timepoints (*i.e.* before and after intervention). First, interaction effects, main group and main time effects were assessed (by combining between-subjects contrast [1 -1 0; 0 1 -1] or [1/3 1/3 1/3], and between-conditions contrast [-1 1] or [1/2 1/2], respectively as fit) with F statistics; followed by *posthoc* pairwise comparisons. *Posthoc* comparisons (totally 9 (within-groups x3, between-groups before x3 and after x3 intervention) were additionally Bonferroni-corrected for multiple testing, resulting in cluster size p-FDR<0.0056 being regarded as significant.

For SBC with brainstem seeds in both hemispheres (e.g. LC and NTS), an F-test (any effect) with both left and right seed was performed first, and, if significance was observed at the additionally multiplicity corrected level (*i.e.* cluster size p-FDR<0.0063 for correlation analyses with baseline factors, and <0.0056 for intervention effects), left and right seeds were also tested separately (T statistics). For all bilateral brainstem seeds (e.g. VTA, PAG, DRN), T statistics were applied directly.

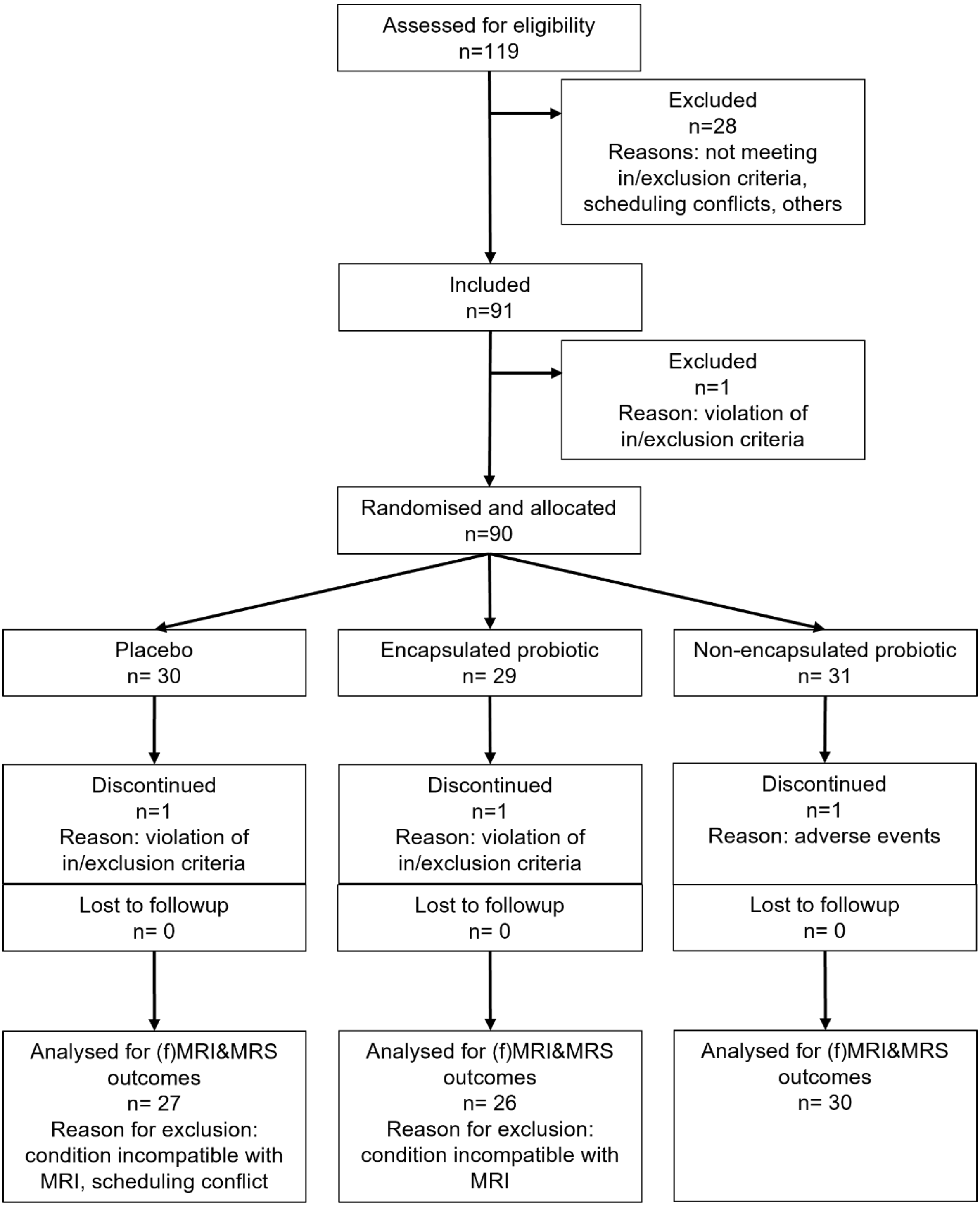


*Supplementary Figure 1. Flow chart (adapted from Rode&Hutchinson et al. 2025)*[1]*. 91 study subjects were recruited, and 90 study subjects were randomised into the three arms: placebo, encapsulated probiotic and non-encapsulated probiotic. After randomisation, in this per protocol study, two subjects were excluded, and one dropped out, leading to a total of 87 study subjects completing the study, whereof n=83 had complete functional magnetic resonance imaging (fMRI) datasets which were analysed in the herein presented work.*

*Supplementary Table 1. Binary brainstem masks obtained*

| Brainstem nucleus | Brainstem region | Hemisphere | Obtained masks | Atlas | Colour in Supplementary Figure 2 |
|---|---|---|---|---|---|
| Subthalamic Nucleus (STN) | Diencephalon (often grouped with midbrain systems) | Paired left/right | Separate left and right | PD25 | blue |
| Substantia Nigra (SN) | Midbrain | Paired left/right | Separate left and right | PD25 | orange |
| Ventral Tegmental Area (VTA) | Midbrain | Mostly midline | One bilateral | Levinson Bari | light blue |
| Periaqueductal Gray (PAG) | Midbrain | Midline | One bilateral | Levinson Bari | yellow |
| Red Nucleus (RN) | Midbrain | Paired left/right | Separate left and right | PD25 | dark red |
| Dorsal Raphe Nucleus (DRN) | Midbrain | Midline | One bilateral | Levinson Bari | red |
| Locus Coeruleus (LC) | Pons | Paired left/right | Separate left and right | Levinson Bari | brown |
| Nucleus Tractus Solitarius (NTS) | Medulla oblongata | Paired left/right | Separate left and right | Levinson Bari | green |

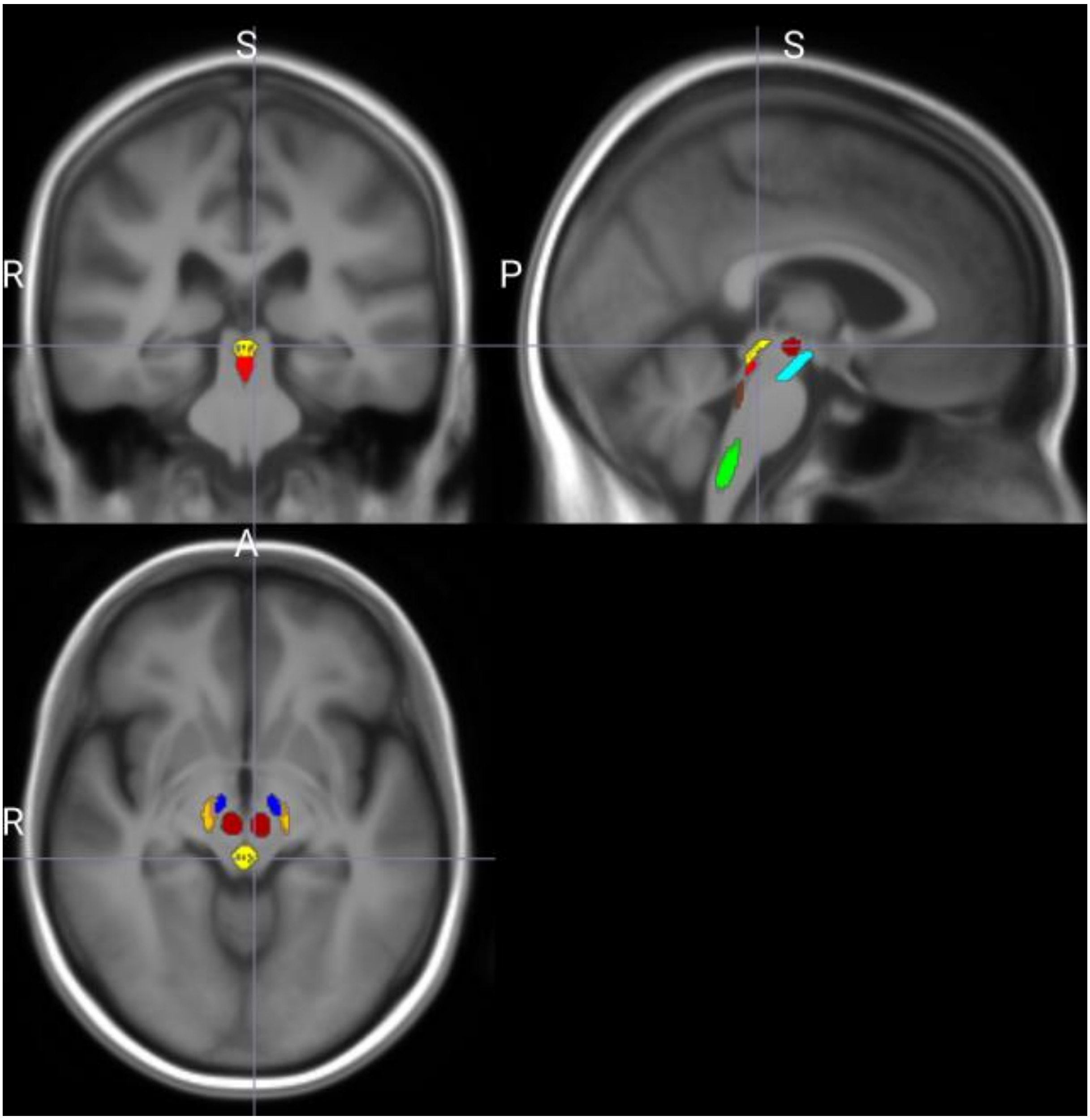


*Supplementary Figure 2. Visualisation of brainstem masks on a mean structural T1-weighted image of all n=83 participants before intervention visualised in the coronal, sagittal and axial plane in Montreal Neurological Institute (MNI) space. Blue shows subthalamic nucleus (STN), orange substantia nigra (SN), light blue ventral tegmental area (VTA), yellow periaqueductal gray (PAG), dark red shows red nucleus (RN), red dorsal raphe nucleus (DRN), brown locus coeruleus (LC), green nucleus tractus solitarius (NTS).*

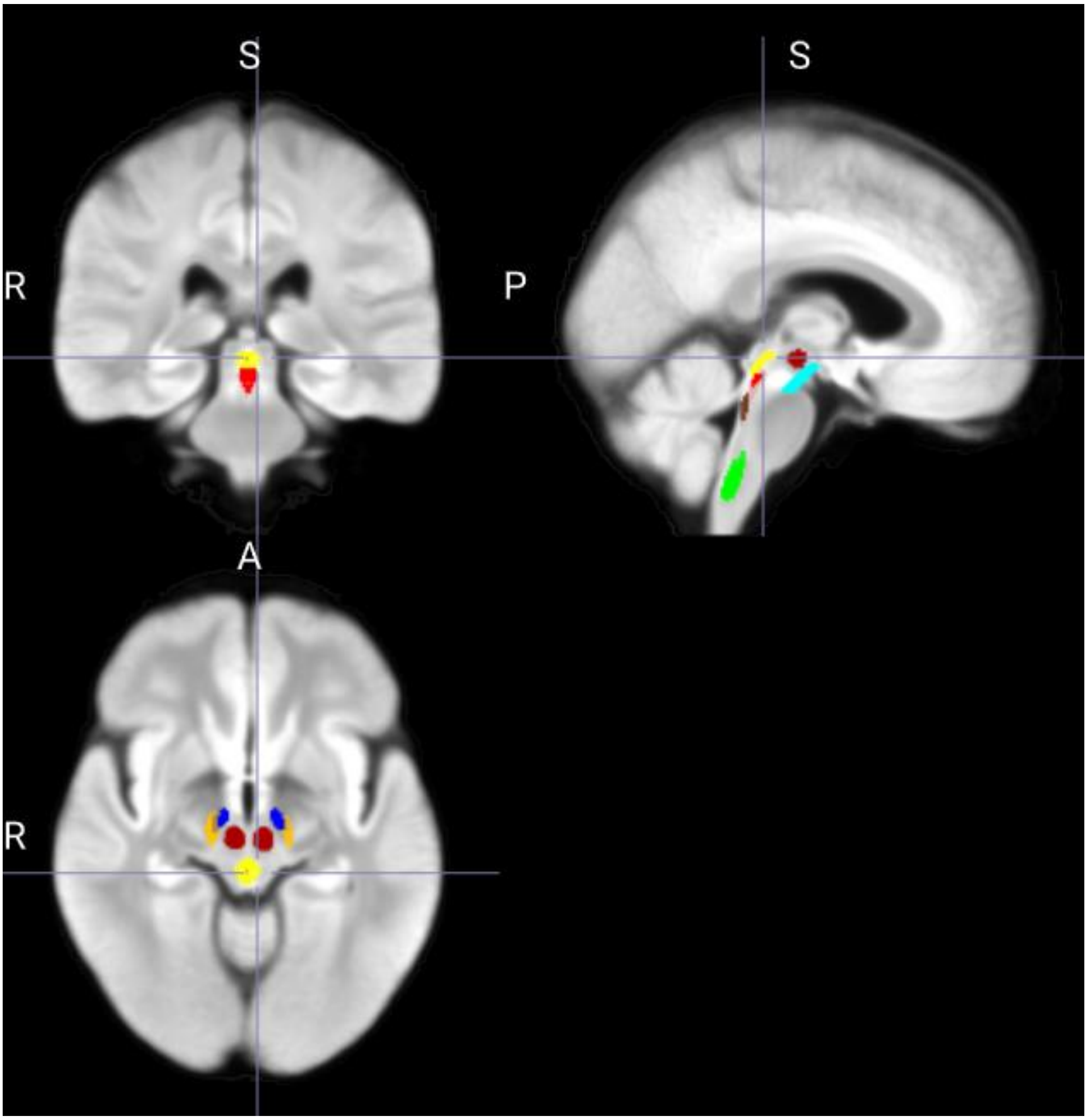


*Supplementary Figure 3. Visualisation of brainstem masks on a mean structural fluid attenuated inversion recovery image of all n=83 participants before intervention visualised in the coronal, sagittal and axial plane in Montreal Neurological Institute (MNI) space. Blue shows subthalamic nucleus (STN), orange substantia nigra (SN), light blue ventral tegmental area (VTA), yellow periaqueductal gray (PAG), dark red shows red nucleus (RN), red dorsal raphe nucleus (DRN), brown locus coeruleus (LC), green nucleus tractus solitarius (NTS).*

*Supplementary Table 2. Manually defined ROI order/groups*

**Thalamocortical relay network**

atlas.Thalamus r

atlas.Thalamus l

**Language network**

atlas.IFG tri r (Inferior Frontal Gyrus, pars triangularis Right)

atlas.IFG tri l (Inferior Frontal Gyrus, pars triangularis Left)

atlas.IFG oper r (Inferior Frontal Gyrus, pars opercularis Right)

atlas.IFG oper l (Inferior Frontal Gyrus, pars opercularis Left)

atlas.aSTG r (Superior Temporal Gyrus, anterior division Right)

atlas.aSTG l (Superior Temporal Gyrus, anterior division Left)

atlas.aMTG r (Middle Temporal Gyrus, anterior division Right)

atlas.aMTG l (Middle Temporal Gyrus, anterior division Left)

atlas.pMTG r (Middle Temporal Gyrus, posterior division Right)

atlas.pMTG l (Middle Temporal Gyrus, posterior division Left)

atlas.aSMG r (Supramarginal Gyrus, anterior division Right)

atlas.aSMG l (Supramarginal Gyrus, anterior division Left)

atlas.AG r (Angular Gyrus Right)

atlas.AG l (Angular Gyrus Left)

atlas.TP r (Temporal Pole Right)

atlas.TP l (Temporal Pole Left)

**Primary visual network**

atlas.ICC r (Intracalcarine Cortex Right)

atlas.ICC l (Intracalcarine Cortex Left)

atlas.SCC r (Supracalcarine Cortex Right)

atlas.SCC l (Supracalcarine Cortex Left)

atlas.OP r (Occipital Pole Right)

atlas.OP l (Occipital Pole Left)

atlas.Cuneal r (Cuneal Cortex Right)

atlas.Cuneal l (Cuneal Cortex Left)

atlas.LG r (Lingual Gyrus Right)

atlas.LG l (Lingual Gyrus Left)

**Ventral visual pathway (ventral stream)**

atlas.sLOC r (Lateral Occipital Cortex, superior division Right)

atlas.sLOC l (Lateral Occipital Cortex, superior division Left)

atlas.iLOC r (Lateral Occipital Cortex, inferior division Right)

atlas.iLOC l (Lateral Occipital Cortex, inferior division Left)

atlas.OFusG r (Occipital Fusiform Gyrus Right)

atlas.OFusG l (Occipital Fusiform Gyrus Left)

atlas.TOFusC r (Temporal Occipital Fusiform Cortex Right)

atlas.TOFusC l (Temporal Occipital Fusiform Cortex Left)

atlas.pTFusC r (Temporal Fusiform Cortex, posterior division Right)

atlas.pTFusC l (Temporal Fusiform Cortex, posterior division Left)

atlas.aTFusC r (Temporal Fusiform Cortex, anterior division Right)

atlas.aTFusC l (Temporal Fusiform Cortex, anterior division Left)

atlas.toITG r (Inferior Temporal Gyrus, temporooccipital part Right)

atlas.toITG l (Inferior Temporal Gyrus, temporooccipital part Left)

atlas.toMTG r (Middle Temporal Gyrus, temporooccipital part Right)

atlas.toMTG l (Middle Temporal Gyrus, temporooccipital part Left)

atlas.pITG r (Inferior Temporal Gyrus, posterior division Right)

atlas.pITG l (Inferior Temporal Gyrus, posterior division Left)

atlas.aITG r (Inferior Temporal Gyrus, anterior division Right)

atlas.aITG l (Inferior Temporal Gyrus, anterior division Left)

**Default mode network**

atlas.MedFC (Frontal Medial Cortex)

atlas.PC (Cingulate Gyrus, posterior division)

atlas.Precuneous (Precuneous Cortex)

atlas.SubCalC (Subcallosal Cortex)

**Auditory network (auditory processing)**

atlas.HG r (Heschl's Gyrus Right)

atlas.HG l (Heschl's Gyrus Left)

atlas.PP r (Planum Polare Right)

atlas.PP l (Planum Polare Left)

atlas.PT r (Planum Temporale Right)

atlas.PT l (Planum Temporale Left)

atlas.pSTG r (Superior Temporal Gyrus, posterior division Right)

atlas.pSTG l (Superior Temporal Gyrus, posterior division Left)

**Executive control**

atlas.SFG r (Superior Frontal Gyrus Right)

atlas.SFG l (Superior Frontal Gyrus Left)

atlas.FP r (Frontal Pole Right)

atlas.FP l (Frontal Pole Left)

**Somatomotor network**

atlas.PreCG r (Precentral Gyrus Right)

atlas.PreCG l (Precentral Gyrus Left)

atlas.PostCG r (Postcentral Gyrus Right)

atlas.PostCG l (Postcentral Gyrus Left)

atlas.SMA r (Juxtapositional Lobule Cortex -formerly Supplementary Motor Cortex- Right)

atlas.SMA L(Juxtapositional Lobule Cortex -formerly Supplementary Motor Cortex- Left)

atlas.CO r (Central Opercular Cortex Right)

atlas.CO l (Central Opercular Cortex Left)

atlas.PO r (Parietal Operculum Cortex Right)

atlas.PO l (Parietal Operculum Cortex Left)

**Dorsal attention network**

atlas.SPL r (Superior Parietal Lobule Right)

atlas.SPL l (Superior Parietal Lobule Left)

atlas.MidFG r (Middle Frontal Gyrus Right)

atlas.MidFG l (Middle Frontal Gyrus Left)

**Salience network**

atlas.IC r (Insular Cortex Right)

atlas.IC l (Insular Cortex Left)

atlas.FO r (Frontal Operculum Cortex Right)

atlas.FO l (Frontal Operculum Cortex Left)

atlas.AC (Cingulate Gyrus, anterior division)

atlas.PaCiG r (Paracingulate Gyrus Right)

atlas.PaCiG l (Paracingulate Gyrus Left)

atlas.pSMG r (Supramarginal Gyrus, posterior division Right)

atlas.pSMG l (Supramarginal Gyrus, posterior division Left)

**Medial temporal network**

atlas.aPaHC r (Parahippocampal Gyrus, anterior division Right)

atlas.aPaHC l (Parahippocampal Gyrus, anterior division Left)

atlas.pPaHC r (Parahippocampal Gyrus, posterior division Right)

atlas.pPaHC l (Parahippocampal Gyrus, posterior division Left)

atlas.Hippocampus r

atlas.Hippocampus l

**Reward/limbic**

atlas.Amygdala r

atlas.Amygdala l

atlas.Accumbens r

atlas.Accumbens l

atlas.FOrb r (Frontal Orbital Cortex Right)

atlas.FOrb l (Frontal Orbital Cortex Left)

**Basal ganglia**

atlas.Caudate r

atlas.Caudate l

atlas.Putamen r

atlas.Putamen l

atlas.Pallidum r

atlas.Pallidum l

**Cerebellum**

atlas.Cereb1 r (Cerebelum Crus1 Right)

atlas.Cereb1 l (Cerebelum Crus1 Left)

atlas.Cereb2 r (Cerebelum Crus2 Right)

atlas.Cereb2 l (Cerebelum Crus2 Left)

atlas.Cereb3 r (Cerebelum 3 Right)

atlas.Cereb3 l (Cerebelum 3 Left)

atlas.Cereb45 r (Cerebelum 4 5 Right)

atlas.Cereb45 l (Cerebelum 4 5 Left)

atlas.Cereb6 r (Cerebelum 6 Right)

atlas.Cereb6 l (Cerebelum 6 Left)

atlas.Cereb7 r (Cerebelum 7b Right)

atlas.Cereb7 l (Cerebelum 7b Left)

atlas.Cereb8 r (Cerebelum 8 Right)

atlas.Cereb8 l (Cerebelum 8 Left)

atlas.Cereb9 r (Cerebelum 9 Right)

atlas.Cereb9 l (Cerebelum 9 Left)

atlas.Cereb10 r (Cerebelum 10 Right)

atlas.Cereb10 l (Cerebelum 10 Left)

atlas.Ver12 (Vermis 1 2)

atlas.Ver3 (Vermis 3)

atlas.Ver45 (Vermis 4 5)

atlas.Ver6 (Vermis 6)

atlas.Ver7 (Vermis 7)

atlas.Ver8 (Vermis 8)

atlas.Ver9 (Vermis 9)

atlas.Ver10 (Vermis 10)

**Brainstem**

STN r (SubthalamicNucleus_R)

STN l (SubthalamicNucleus_L)

SN r (SubstantiaNigra_R)

SN l (SubstantiaNigra_L)

VTA (VentralTegmentalArea)

PAG (PeriaqueductalGray)

RN r (RedNucleus_R)

RN l (RedNucleus_L)

DRN (DorsalRapheNucleus)

LC r (LocusCoeruleus_R)

LC l (LocusCoeruleus_L)

NTS r (NucleusTractusSolitarius_R)

NTS l (NucleusTractusSolitarius_L)

*Supplementary Table 3. Within-brainstem (first below presented cluster 57/120) and region-based brainstem-to-cortex/subcortex/cerebellum functional connectivity with brainstem seeds at baseline in all n=83 participants, RRC. Clusters demonstrating within-cortex and cortex-to-cerebellum functional connectivity are not presented since they are out of scope of the herein presented work.*

| Analysis Unit | Statistic | p-unc | p-FDR |
|---|---|---|---|
| Cluster 57/120 | F(3,80) = 128.97 | 0.000000 | 0.000000 |
| Connection NucleusTractusSolitarius_R - NucleusTractusSolitarius_L | T(82) = 12.32 | 0.000000 | 0.000000 |
| Connection LocusCoeruleus_R - LocusCoeruleus_L | T(82) = 10.31 | 0.000000 | 0.000000 |
| Connection SubstantiaNigra_R - VentralTegmentalArea | T(82) = 9.84 | 0.000000 | 0.000000 |
| Connection PeriaqueductalGray - DorsalRapheNucleus | T(82) = 8.47 | 0.000000 | 0.000000 |
| Connection DorsalRapheNucleus - LocusCoeruleus_R | T(82) = 7.81 | 0.000000 | 0.000000 |
| Connection DorsalRapheNucleus - LocusCoeruleus_L | T(82) = 6.98 | 0.000000 | 0.000000 |
| Connection RedNucleus_R - RedNucleus_L | T(82) = 6.15 | 0.000000 | 0.000004 |
| Connection VentralTegmentalArea - PeriaqueductalGray | T(82) = 5.26 | 0.000001 | 0.000018 |
| Connection RedNucleus_R - DorsalRapheNucleus | T(82) = -3.06 | 0.003009 | 0.015937 |
| Connection PeriaqueductalGray - LocusCoeruleus_L | T(82) = 2.91 | 0.004585 | 0.026229 |
| Connection VentralTegmentalArea - LocusCoeruleus_R | T(82) = 2.75 | 0.007368 | 0.027017 |
| Connection SubThalamicNucleus_R - SubstantiaNigra_R | T(82) = 3.91 | 0.000192 | 0.027404 |
| Connection SubstantiaNigra_R - SubstantiaNigra_L | T(82) = 2.91 | 0.004677 | 0.037157 |
| Connection SubstantiaNigra_L - VentralTegmentalArea | T(82) = 3.37 | 0.001159 | 0.043126 |
| Connection LocusCoeruleus_L - NucleusTractusSolitarius_R | T(82) = 2.53 | 0.013480 | 0.045896 |
| Connection PeriaqueductalGray - LocusCoeruleus_R | T(82) = 2.47 | 0.015494 | 0.065165 |
| Connection DorsalRapheNucleus - NucleusTractusSolitarius_L | T(82) = 2.19 | 0.031091 | 0.069470 |
| Connection PeriaqueductalGray - RedNucleus_L | T(82) = 2.25 | 0.027060 | 0.104585 |
| Connection LocusCoeruleus_R - NucleusTractusSolitarius_R | T(82) = 2.18 | 0.031773 | 0.112482 |
| Connection PeriaqueductalGray - NucleusTractusSolitarius_L | T(82) = 2.20 | 0.030729 | 0.112673 |
| Connection SubstantiaNigra_R - PeriaqueductalGray | T(82) = 2.15 | 0.034714 | 0.171176 |
| Connection SubstantiaNigra_R - RedNucleus_R | T(82) = 2.03 | 0.045962 | 0.182573 |
| Connection SubThalamicNucleus_R - PeriaqueductalGray | T(82) = 2.67 | 0.009031 | 0.340379 |
| Connection SubThalamicNucleus_R - DorsalRapheNucleus | T(82) = 2.15 | 0.034167 | 0.444176 |

Cluster 63/120    F(3,80) = 103.83    0.000000    0.000000

Connection atlas.Ver3 (Vermis 3) - PeriaqueductalGray    T(82) = 16.44    0.000000    0.000000

Connection atlas.Ver12 (Vermis 1 2) - DorsalRapheNucleus    T(82) = 13.89    0.000000    0.000000

Connection atlas.Ver12 (Vermis 1 2) - LocusCoeruleus_L    T(82) = 10.11    0.000000    0.000000

Connection atlas.Ver12 (Vermis 1 2) - LocusCoeruleus_R    T(82) = 10.10    0.000000    0.000000

Connection atlas.Cereb45 l (Cerebelum 4 5 Left) - PeriaqueductalGray    T(82) = 8.63    0.000000    0.000000

Connection atlas.Cereb8 r (Cerebelum 8 Right) - NucleusTractusSolitarius_R    T(82) = 8.39    0.000000    0.000000

Connection atlas.Cereb45 r (Cerebelum 4 5 Right) - PeriaqueductalGray    T(82) = 8.27    0.000000    0.000000

Connection atlas.Cereb8 l (Cerebelum 8 Left) - NucleusTractusSolitarius_R    T(82) = 8.12    0.000000    0.000000

Connection atlas.Cereb8 l (Cerebelum 8 Left) - NucleusTractusSolitarius_L    T(82) = 7.51    0.000000    0.000000

Connection atlas.Cereb9 r (Cerebelum 9 Right) - NucleusTractusSolitarius_R    T(82) = 7.56    0.000000    0.000000

Connection atlas.Cereb6 l (Cerebelum 6 Left) - DorsalRapheNucleus    T(82) = 7.33    0.000000    0.000000

Connection atlas.Cereb45 l (Cerebelum 4 5 Left) - DorsalRapheNucleus    T(82) = 7.37    0.000000    0.000000

Connection atlas.Cereb6 r (Cerebelum 6 Right) - LocusCoeruleus_L    T(82) = 7.23    0.000000    0.000000

Connection atlas.Cereb9 r (Cerebelum 9 Right) - NucleusTractusSolitarius_L    T(82) = 7.29    0.000000    0.000000

Connection atlas.Ver45 (Vermis 4 5) - DorsalRapheNucleus    T(82) = 6.79    0.000000    0.000000

Connection atlas.Cereb8 r (Cerebelum 8 Right) - NucleusTractusSolitarius_L    T(82) = 6.67    0.000000    0.000000

Connection atlas.Cereb6 l (Cerebelum 6 Left) - LocusCoeruleus_L    T(82) = 6.44    0.000000    0.000000

Connection atlas.Cereb6 l (Cerebelum 6 Left) - NucleusTractusSolitarius_R    T(82) = 6.32    0.000000    0.000000

Connection atlas.Ver45 (Vermis 4 5) - PeriaqueductalGray    T(82) = 6.43    0.000000    0.000000

Connection atlas.Cereb9 l (Cerebelum 9 Left) - NucleusTractusSolitarius_R    T(82) = 6.34    0.000000    0.000000

Connection atlas.Ver8 (Vermis 8) - LocusCoeruleus_L    T(82) = 6.15    0.000000    0.000000

Connection atlas.Cereb9 l (Cerebelum 9 Left) - NucleusTractusSolitarius_L    T(82) = 6.12    0.000000    0.000000

Connection atlas.Cereb6 r (Cerebelum 6 Right) - DorsalRapheNucleus  T(82) = 5.93  0.000000  0.000000

Connection atlas.Cereb8 r (Cerebelum 8 Right) - LocusCoeruleus_L  T(82) = 6.00  0.000000  0.000000

Connection atlas.Ver6 (Vermis 6) - VentralTegmentalArea  T(82) = 5.86  0.000000  0.000001

Connection atlas.Cereb8 l (Cerebelum 8 Left) - LocusCoeruleus_L  T(82) = 5.75  0.000000  0.000001

Connection atlas.Cereb45 l (Cerebelum 4 5 Left) - LocusCoeruleus_L  T(82) = 5.58  0.000000  0.000001

Connection atlas.Cereb7 l (Cerebelum 7b Left) - NucleusTractusSolitarius_R  T(82) = 5.60  0.000000  0.000001

Connection atlas.Cereb6 l (Cerebelum 6 Left) - LocusCoeruleus_R  T(82) = 5.37  0.000001  0.000001

Connection atlas.Cereb45 l (Cerebelum 4 5 Left) - LocusCoeruleus_R  T(82) = 5.47  0.000000  0.000001

Connection atlas.Ver6 (Vermis 6) - PeriaqueductalGray  T(82) = 5.48  0.000000  0.000002

Connection atlas.Cereb45 r (Cerebelum 4 5 Right) - LocusCoeruleus_L  T(82) = 5.27  0.000001  0.000004

Connection atlas.Cereb6 r (Cerebelum 6 Right) - LocusCoeruleus_R  T(82) = 4.87  0.000005  0.000013

Connection atlas.Cereb7 l (Cerebelum 7b Left) - LocusCoeruleus_L  T(82) = 4.99  0.000003  0.000013

Connection atlas.Ver3 (Vermis 3) - VentralTegmentalArea  T(82) = 5.26  0.000001  0.000014

Connection atlas.Cereb6 l (Cerebelum 6 Left) - PeriaqueductalGray  T(82) = 4.65  0.000012  0.000022

Connection atlas.Ver6 (Vermis 6) - DorsalRapheNucleus  T(82) = 4.86  0.000006  0.000024

Connection atlas.Cereb45 l (Cerebelum 4 5 Left) - NucleusTractusSolitarius_L  T(82) = 4.72  0.000010  0.000025

Connection atlas.Cereb6 l (Cerebelum 6 Left) - NucleusTractusSolitarius_L  T(82) = 4.53  0.000020  0.000034

Connection atlas.Ver8 (Vermis 8) - NucleusTractusSolitarius_R  T(82) = 4.78  0.000008  0.000038

Connection atlas.Cereb6 r (Cerebelum 6 Right) - NucleusTractusSolitarius_R  T(82) = 4.56  0.000018  0.000039

Connection atlas.Cereb7 r (Cerebelum 7b Right) - LocusCoeruleus_L  T(82) = 4.63  0.000013  0.000043

Connection atlas.Cereb45 l (Cerebelum 4 5 Left) - VentralTegmentalArea  T(82) = 4.52  0.000021  0.000051

Connection atlas.Cereb10 l (Cerebelum 10 Left) - DorsalRapheNucleus  T(82) = 4.81  0.000007  0.000061

Connection atlas.Ver8 (Vermis 8) - NucleusTractusSolitarius_L  T(82) = 4.60  0.000015  0.000071

Connection atlas.Ver7 (Vermis 7) - LocusCoeruleus_R  T(82) = 4.52  0.000020  0.000088

Connection atlas.Cereb45 r (Cerebelum 4 5 Right) - LocusCoeruleus_R T(82) = 4.38 0.000035 0.000088

Connection atlas.Cereb45 r (Cerebelum 4 5 Right) - VentralTegmentalArea T(82) = 4.35 0.000039 0.000095

Connection atlas.Cereb7 l (Cerebelum 7b Left) - NucleusTractusSolitarius_L T(82) = 4.41 0.000032 0.000096

Connection atlas.Cereb6 r (Cerebelum 6 Right) - PeriaqueductalGray T(82) = 4.20 0.000068 0.000137

Connection atlas.Cereb3 l (Cerebelum 3 Left) - PeriaqueductalGray T(82) = 4.26 0.000054 0.000149

Connection atlas.Cereb3 l (Cerebelum 3 Left) - LocusCoeruleus_R T(82) = 4.22 0.000063 0.000170

Connection atlas.Cereb2 l (Cerebelum Crus2 Left) - LocusCoeruleus_L T(82) = 4.11 0.000093 0.000185

Connection atlas.Cereb8 r (Cerebelum 8 Right) - LocusCoeruleus_R T(82) = 4.16 0.000079 0.000190

Connection atlas.Cereb7 r (Cerebelum 7b Right) - LocusCoeruleus_R T(82) = 4.10 0.000095 0.000235

Connection atlas.Cereb6 r (Cerebelum 6 Right) - NucleusTractusSolitarius_L T(82) = 3.99 0.000141 0.000265

Connection atlas.Cereb10 r (Cerebelum 10 Right) - NucleusTractusSolitarius_R T(82) = 4.27 0.000051 0.000272

Connection atlas.Cereb7 r (Cerebelum 7b Right) - NucleusTractusSolitarius_R T(82) = 4.03 0.000126 0.000301

Connection atlas.Ver6 (Vermis 6) - NucleusTractusSolitarius_L T(82) = 4.07 0.000108 0.000336

Connection atlas.Cereb45 r (Cerebelum 4 5 Right) - NucleusTractusSolitarius_R T(82) = 3.97 0.000152 0.000345

Connection atlas.Cereb1 l (Cerebelum Crus1 Left) - NucleusTractusSolitarius_R T(82) = 3.92 0.000182 0.000383

Connection atlas.Cereb45 l (Cerebelum 4 5 Left) - NucleusTractusSolitarius_R T(82) = 3.87 0.000219 0.000402

Connection atlas.Cereb8 l (Cerebelum 8 Left) - LocusCoeruleus_R T(82) = 3.93 0.000174 0.000414

Connection atlas.Cereb9 r (Cerebelum 9 Right) - LocusCoeruleus_L T(82) = 3.90 0.000198 0.000480

Connection atlas.Cereb45 r (Cerebelum 4 5 Right) - DorsalRapheNucleus T(82) = 3.85 0.000229 0.000489

Connection atlas.Cereb10 l (Cerebelum 10 Left) - NucleusTractusSolitarius_R T(82) = 4.10 0.000097 0.000495

Connection atlas.Cereb3 r (Cerebelum 3 Right) - PeriaqueductalGray T(82) = 3.89 0.000202 0.000556

Connection atlas.Cereb1 l (Cerebelum Crus1 Left) - LocusCoeruleus_L T(82) = 3.79 0.000284 0.000564

Connection atlas.Cereb7 r (Cerebelum 7b Right) - NucleusTractusSolitarius_L T(82) = 3.82 0.000257 0.000603

Connection atlas.Cereb10 r (Cerebelum 10 Right) - NucleusTractusSolitarius_L T(82) = 3.99 0.000145 0.000626

Connection atlas.Cereb1 l (Cerebelum Crus1 Left) - DorsalRapheNucleus T(82) = 3.71 0.000376 0.000737

Connection atlas.Ver6 (Vermis 6) - LocusCoeruleus_L T(82) = 3.80 0.000275 0.000741

Connection atlas.Ver6 (Vermis 6) - NucleusTractusSolitarius_R T(82) = 3.79 0.000286 0.000756

Connection atlas.Cereb45 r (Cerebelum 4 5 Right) - NucleusTractusSolitarius_L T(82) = 3.66 0.000445 0.000908

Connection atlas.Cereb8 l (Cerebelum 8 Left) - DorsalRapheNucleus T(82) = 3.63 0.000487 0.001088

Connection atlas.Ver7 (Vermis 7) - LocusCoeruleus_L T(82) = 3.66 0.000447 0.001559

Connection atlas.Ver9 (Vermis 9) - NucleusTractusSolitarius_R T(82) = 3.73 0.000352 0.001571

Connection atlas.Cereb2 l (Cerebelum Crus2 Left) - DorsalRapheNucleus T(82) = 3.44 0.000908 0.001686

Connection atlas.Cereb9 r (Cerebelum 9 Right) - DorsalRapheNucleus T(82) = 3.47 0.000833 0.001860

Connection atlas.Ver45 (Vermis 4 5) - NucleusTractusSolitarius_L T(82) = 3.45 0.000889 0.001957

Connection atlas.Cereb9 r (Cerebelum 9 Right) - PeriaqueductalGray T(82) = 3.37 0.001156 0.002500

Connection atlas.Ver9 (Vermis 9) - LocusCoeruleus_L T(82) = 3.52 0.000713 0.002684

Connection atlas.Ver10 (Vermis 10) - NucleusTractusSolitarius_L T(82) = 3.57 0.000599 0.002814

Connection atlas.Cereb9 l (Cerebelum 9 Left) - LocusCoeruleus_L T(82) = 3.44 0.000916 0.002914

Connection atlas.Ver7 (Vermis 7) - VentralTegmentalArea T(82) = 3.43 0.000935 0.003022

Connection atlas.Cereb1 l (Cerebelum Crus1 Left) - NucleusTractusSolitarius_L T(82) = 3.23 0.001788 0.003044

Connection atlas.Cereb2 r (Cerebelum Crus2 Right) - NucleusTractusSolitarius_L T(82) = 3.21 0.001917 0.003045

Connection atlas.Cereb10 r (Cerebelum 10 Right) - DorsalRapheNucleus T(82) = 3.42 0.000964 0.003063

Connection atlas.Cereb9 r (Cerebelum 9 Right) - VentralTegmentalArea T(82) = 3.24 0.001715 0.003407

Connection atlas.Ver45 (Vermis 4 5) - VentralTegmentalArea T(82) = 3.26 0.001636 0.003493

Connection atlas.Cereb7 l (Cerebelum 7b Left) - DorsalRapheNucleus T(82) = 3.21 0.001923 0.003873

Connection atlas.Cereb10 l (Cerebelum 10 Left) - LocusCoeruleus_R T(82) = 3.36 0.001192 0.004158

Connection atlas.Cereb45 r (Cerebelum 4 5 Right) - SubstantiaNigra_R T(82) = 3.12 0.002469 0.004586

Connection atlas.Ver6 (Vermis 6) - LocusCoeruleus_R T(82) = 3.17 0.002129 0.004683

Connection atlas.Cereb2 r (Cerebelum Crus2 Right) - VentralTegmentalArea T(82) = 3.05 0.003058 0.004703

Connection atlas.Cereb7 r (Cerebelum 7b Right) - DorsalRapheNucleus T(82) = 3.16 0.002205 0.004707

Connection atlas.Ver12 (Vermis 1 2) - PeriaqueductalGray T(82) = 3.44 0.000909 0.005418

Connection atlas.Ver45 (Vermis 4 5) - LocusCoeruleus_R T(82) = 3.08 0.002805 0.005814

Connection atlas.Ver3 (Vermis 3) - SubstantiaNigra_R T(82) = 3.30 0.001445 0.005905

Connection atlas.Ver45 (Vermis 4 5) - LocusCoeruleus_L T(82) = 3.06 0.002972 0.006070

Connection atlas.Ver7 (Vermis 7) - DorsalRapheNucleus T(82) = 3.15 0.002273 0.006501

Connection atlas.Ver6 (Vermis 6) - RedNucleus_L T(82) = 3.04 0.003137 0.006597

Connection atlas.Cereb3 r (Cerebelum 3 Right) - LocusCoeruleus_L T(82) = 3.01 0.003422 0.006702

Connection atlas.Cereb9 r (Cerebelum 9 Right) - LocusCoeruleus_R T(82) = 2.97 0.003941 0.006872

Connection atlas.Cereb10 l (Cerebelum 10 Left) - NucleusTractusSolitarius_L T(82) = 3.17 0.002141 0.007290

Connection atlas.Cereb8 r (Cerebelum 8 Right) - DorsalRapheNucleus T(82) = 2.97 0.003905 0.007866

Connection atlas.Cereb2 l (Cerebelum Crus2 Left) - NucleusTractusSolitarius_L T(82) = 2.89 0.004899 0.008146

Connection atlas.Cereb1 r (Cerebelum Crus1 Right) - DorsalRapheNucleus T(82) = 2.85 0.005451 0.008382

Connection atlas.Cereb45 r (Cerebelum 4 5 Right) - RedNucleus_L T(82) = 2.84 0.005709 0.009605

Connection atlas.Ver7 (Vermis 7) - NucleusTractusSolitarius_L T(82) = 2.99 0.003697 0.009653

Connection atlas.Cereb3 r (Cerebelum 3 Right) - LocusCoeruleus_R T(82) = 2.82 0.005998 0.011591

Connection atlas.Cereb9 l (Cerebelum 9 Left) - VentralTegmentalArea T(82) = 2.93 0.004363 0.011999

Connection atlas.Cereb7 l (Cerebelum 7b Left) - LocusCoeruleus_R T(82) = 2.79 0.006487 0.012149

Connection atlas.Cereb10 r (Cerebelum 10 Right) - LocusCoeruleus_R T(82) = 2.90 0.004854 0.012178

Connection atlas.Cereb9 l (Cerebelum 9 Left) - PeriaqueductalGray T(82) = 2.87 0.005273 0.013710

Connection atlas.Cereb2 l (Cerebelum Crus2 Left) - NucleusTractusSolitarius_R T(82) = 2.66 0.009418 0.015132

Connection atlas.Cereb2 r (Cerebelum Crus2 Right) - SubstantiaNigra_R T(82) = 2.59 0.011237 0.016565

Connection atlas.Cereb2 r (Cerebelum Crus2 Right) - NucleusTractusSolitarius_R T(82) = 2.53 0.013225 0.019103

Connection atlas.Ver6 (Vermis 6) - RedNucleus_R T(82) = 2.62 0.010477 0.020808

Connection atlas.Cereb1 r (Cerebelum Crus1 Right) - NucleusTractusSolitarius_L T(82) = 2.44 0.016651 0.023345

Connection atlas.Ver10 (Vermis 10) - NucleusTractusSolitarius_R T(82) = 2.59 0.011303 0.023770

Connection atlas.Ver3 (Vermis 3) - RedNucleus_R T(82) = 2.72 0.008028 0.023918

Connection atlas.Cereb10 r (Cerebelum 10 Right) - LocusCoeruleus_L T(82) = 2.59 0.011398 0.024696

Connection atlas.Cereb9 l (Cerebelum 9 Left) - LocusCoeruleus_R T(82) = 2.62 0.010565 0.024748

Connection atlas.Cereb1 r (Cerebelum Crus1 Right) - LocusCoeruleus_L T(82) = 2.42 0.017885 0.024831

Connection atlas.Cereb9 l (Cerebelum 9 Left) - DorsalRapheNucleus T(82) = 2.60 0.011165 0.025343

Connection atlas.Ver9 (Vermis 9) - NucleusTractusSolitarius_L T(82) = 2.64 0.009864 0.026613

Connection atlas.Cereb2 r (Cerebelum Crus2 Right) - LocusCoeruleus_L T(82) = 2.38 0.019446 0.027807

Connection atlas.Cereb10 l (Cerebelum 10 Left) - PeriaqueductalGray T(82) = 2.65 0.009670 0.028222

Connection atlas.Ver7 (Vermis 7) - PeriaqueductalGray T(82) = 2.56 0.012223 0.029625

Connection atlas.Ver7 (Vermis 7) - NucleusTractusSolitarius_R T(82) = 2.53 0.013483 0.031098

Connection atlas.Cereb9 r (Cerebelum 9 Right) - RedNucleus_R T(82) = 2.35 0.021007 0.032301

Connection atlas.Cereb2 r (Cerebelum Crus2 Right) - RedNucleus_R T(82) = 2.31 0.023193 0.032515

Connection atlas.Ver7 (Vermis 7) - SubstantiaNigra_R T(82) = 2.49 0.014693 0.033351

Connection atlas.Ver3 (Vermis 3) - LocusCoeruleus_R T(82) = 2.45 0.016229 0.040014

Connection atlas.Ver8 (Vermis 8) - LocusCoeruleus_R T(82) = 2.45 0.016517 0.042178

Connection atlas.Cereb45 l (Cerebelum 4 5 Left) - RedNucleus_L T(82) = 2.18 0.032255 0.043107

Connection atlas.Cereb2 l (Cerebelum Crus2 Left) - RedNucleus_L T(82) = 2.23 0.028498 0.043354

Connection atlas.Ver9 (Vermis 9) - DorsalRapheNucleus T(82) = 2.40 0.018743 0.043938

Connection atlas.Cereb45 r (Cerebelum 4 5 Right) - SubThalamicNucleus_R T(82) = 2.19 0.031202 0.044177

Connection atlas.Ver3 (Vermis 3) - NucleusTractusSolitarius_L T(82) = 2.37 0.019909 0.046671

Connection atlas.Cereb2 r (Cerebelum Crus2 Right) - DorsalRapheNucleus T(82) = 2.15 0.034457 0.046927

Connection atlas.Cereb6 r (Cerebelum 6 Right) - VentralTegmentalArea T(82) = 2.12 0.036963 0.047619

Connection atlas.Cereb2 r (Cerebelum Crus2 Right) - LocusCoeruleus_R T(82) = 2.13 0.036145 0.048761

Connection atlas.Cereb3 l (Cerebelum 3 Left) - LocusCoeruleus_L T(82) = 2.19 0.031473 0.050007

Connection atlas.Cereb7 r (Cerebelum 7b Right) - VentralTegmentalArea T(82) = 2.16 0.033503 0.054442

Connection atlas.Cereb1 l (Cerebelum Crus1 Left) - LocusCoeruleus_R T(82) = 2.10 0.039067 0.054771

Connection atlas.Cereb45 r (Cerebelum 4 5 Right) - RedNucleus_R T(82) = 2.08 0.040381 0.056063

Connection atlas.Cereb3 l (Cerebelum 3 Left) - RedNucleus_R T(82) = 2.11 0.037542 0.058353

Connection atlas.Ver12 (Vermis 1 2) - SubThalamicNucleus_R T(82) = 2.50 0.014392 0.058803

Connection atlas.Cereb3 r (Cerebelum 3 Right) - NucleusTractusSolitarius_L T(82) = 2.09 0.039353 0.060394

Connection atlas.Cereb9 l (Cerebelum 9 Left) - SubstantiaNigra_R T(82) = 2.11 0.037927 0.071803

Connection atlas.Ver3 (Vermis 3) - RedNucleus_L T(82) = 2.05 0.043471 0.092782

Connection atlas.Ver12 (Vermis 1 2) - VentralTegmentalArea T(82) = 2.06 0.042088 0.136785

Cluster 82/120 F(3,80) = 44.92 0.000000 0.000000

Connection atlas.aPaHC l (Parahippocampal Gyrus, anterior division Left) - DorsalRapheNucleus T(82) = 8.37 0.000000 0.000000

Connection atlas.pPaHC l (Parahippocampal Gyrus, posterior division Left) - DorsalRapheNucleus T(82) = 8.18 0.000000 0.000000

Connection atlas.pPaHC r (Parahippocampal Gyrus, posterior division Right) - DorsalRapheNucleus T(82) = 8.02 0.000000 0.000000

Connection atlas.aPaHC r (Parahippocampal Gyrus, anterior division Right) - DorsalRapheNucleus T(82) = 7.17 0.000000 0.000000

Connection atlas.Hippocampus l - DorsalRapheNucleus T(82) = 5.97 0.000000 0.000000

Connection atlas.aPaHC l (Parahippocampal Gyrus, anterior division Left) - LocusCoeruleus_L T(82) = 5.86 0.000000 0.000000

Connection atlas.aPaHC r (Parahippocampal Gyrus, anterior division Right) - LocusCoeruleus_L T(82) = 5.14 0.000002 0.000007

Connection atlas.pPaHC l (Parahippocampal Gyrus, posterior division Left) - LocusCoeruleus_L T(82) = 4.99 0.000003 0.000009

Connection atlas.aPaHC l (Parahippocampal Gyrus, anterior division Left) - LocusCoeruleus_R T(82) = 4.81 0.000007 0.000019

Connection atlas.pPaHC r (Parahippocampal Gyrus, posterior division Right) - LocusCoeruleus_L T(82) = 4.47 0.000025 0.000071

Connection atlas.pPaHC l (Parahippocampal Gyrus, posterior division Left) - LocusCoeruleus_R T(82) = 4.43 0.000029 0.000073

Connection atlas.aPaHC r (Parahippocampal Gyrus, anterior division Right) - LocusCoeruleus_R T(82) = 4.47 0.000025 0.000079

Connection atlas.Hippocampus r - DorsalRapheNucleus T(82) = 4.38 0.000035 0.000079

Connection atlas.pPaHC l (Parahippocampal Gyrus, posterior division Left) - PeriaqueductalGray T(82) = 4.18 0.000072 0.000170

Connection atlas.pPaHC r (Parahippocampal Gyrus, posterior division Right) - PeriaqueductalGray T(82) = 4.14 0.000083 0.000215

Connection atlas.Hippocampus r - VentralTegmentalArea T(82) = 3.73 0.000354 0.000657

Connection atlas.aPaHC r (Parahippocampal Gyrus, anterior division Right) - PeriaqueductalGray T(82) = 3.62 0.000502 0.001216

Connection atlas.Hippocampus l - LocusCoeruleus_L T(82) = 3.18 0.002053 0.004078

Connection atlas.Hippocampus l - PeriaqueductalGray T(82) = 3.17 0.002140 0.004190

Connection atlas.pPaHC r (Parahippocampal Gyrus, posterior division Right) - LocusCoeruleus_R T(82) = 2.88 0.005004 0.008945

Connection atlas.Hippocampus l - LocusCoeruleus_R T(82) = 2.87 0.005147 0.009087

Connection atlas.Hippocampus r - LocusCoeruleus_L T(82) = 2.84 0.005620 0.009606

Connection atlas.pPaHC l (Parahippocampal Gyrus, posterior division Left) - VentralTegmentalArea T(82) = 2.80 0.006415 0.010424

Connection atlas.Hippocampus r - PeriaqueductalGray T(82) = 2.77 0.006907 0.011486

Connection atlas.pPaHC r (Parahippocampal Gyrus, posterior division Right) - SubstantiaNigra_L T(82) = 2.54 0.013052 0.022219

Connection atlas.Hippocampus r - LocusCoeruleus_R T(82) = 2.27 0.026111 0.041032

Connection atlas.pPaHC l (Parahippocampal Gyrus, posterior division Left) - SubstantiaNigra_R T(82) = 2.04 0.044929 0.060612

Connection atlas.Hippocampus l - VentralTegmentalArea T(82) = 2.02 0.046387 0.069097

Connection atlas.aPaHC r (Parahippocampal Gyrus, anterior division Right) - RedNucleus_R T(82) = 2.01 0.048138 0.078224

Connection atlas.aPaHC l (Parahippocampal Gyrus, anterior division Left) - PeriaqueductalGray T(82) = 1.99 0.049459 0.088407

Cluster 86/120 F(3,80) = 35.38 0.000000 0.000000

Connection atlas.pTFusC l (Temporal Fusiform Cortex, posterior division Left) - DorsalRapheNucleus T(82) = 8.18 0.000000 0.000000

Connection atlas.pTFusC l (Temporal Fusiform Cortex, posterior division Left) - LocusCoeruleus_R T(82) = 6.38 0.000000 0.000000

Connection atlas.pTFusC l (Temporal Fusiform Cortex, posterior division Left) - LocusCoeruleus_L T(82) = 5.67 0.000000 0.000001

Connection atlas.pITG l (Inferior Temporal Gyrus, posterior division Left) - RedNucleus_R T(82) = 4.79 0.000007 0.000020

Connection atlas.aTFusC r (Temporal Fusiform Cortex, anterior division Right) - LocusCoeruleus_L T(82) = 4.86 0.000006 0.000043

Connection atlas.aTFusC r (Temporal Fusiform Cortex, anterior division Right) - DorsalRapheNucleus T(82) = 4.85 0.000006 0.000043

Connection atlas.toITG l (Inferior Temporal Gyrus, temporooccipital part Left) - RedNucleus_R T(82) = 4.45 0.000027 0.000086

Connection atlas.pITG l (Inferior Temporal Gyrus, posterior division Left) - NucleusTractusSolitarius_R T(82) = -4.28 0.000051 0.000132

Connection atlas.aTFusC l (Temporal Fusiform Cortex, anterior division Left) - LocusCoeruleus_L T(82) = 4.39 0.000034 0.000200

Connection atlas.pITG r (Inferior Temporal Gyrus, posterior division Right) - NucleusTractusSolitarius_R T(82) = -4.04 0.000120 0.000342

Connection atlas.TOFusC l (Temporal Occipital Fusiform Cortex Left) - DorsalRapheNucleus T(82) = 3.75 0.000324 0.000634

Connection atlas.TOFusC r (Temporal Occipital Fusiform Cortex Right) - SubstantiaNigra_L T(82) = 3.60 0.000544 0.001096

Connection atlas.TOFusC r (Temporal Occipital Fusiform Cortex Right) - DorsalRapheNucleus T(82) = 3.47 0.000828 0.001621

Connection atlas.pTFusC r (Temporal Fusiform Cortex, posterior division Right) - DorsalRapheNucleus T(82) = 3.55 0.000652 0.001694

Connection atlas.aTFusC l (Temporal Fusiform Cortex, anterior division Left) - LocusCoeruleus_R T(82) = 3.65 0.000453 0.002233

Connection atlas.toMTG r (Middle Temporal Gyrus, temporooccipital part Right) - SubstantiaNigra_L T(82) = 3.28 0.001508 0.002695

Connection atlas.aITG l (Inferior Temporal Gyrus, anterior division Left) - SubstantiaNigra_R T(82) = 3.34 0.001244 0.002868

Connection atlas.aITG r (Inferior Temporal Gyrus, anterior division Right) - RedNucleus_R T(82) = 3.41 0.001011 0.002950

Connection atlas.aTFusC l (Temporal Fusiform Cortex, anterior division Left) - DorsalRapheNucleus T(82) = 3.34 0.001278 0.005221

Connection atlas.iLOC r (Lateral Occipital Cortex, inferior division Right) - NucleusTractusSolitarius_L T(82) = -3.01 0.003474 0.006629

Connection atlas.pTFusC r (Temporal Fusiform Cortex, posterior division Right) - RedNucleus_R T(82) = 2.97 0.003919 0.008492

Connection atlas.pTFusC r (Temporal Fusiform Cortex, posterior division Right) - SubstantiaNigra_L T(82) = 2.92 0.004505 0.009614

Connection atlas.iLOC l (Lateral Occipital Cortex, inferior division Left) - SubstantiaNigra_L T(82) = 2.77 0.006857 0.012256

Connection atlas.iLOC r (Lateral Occipital Cortex, inferior division Right) - VentralTegmentalArea T(82) = -2.77 0.006940 0.012724

Connection atlas.pTFusC l (Temporal Fusiform Cortex, posterior division Left) - SubThalamicNucleus_R T(82) = 2.61 0.010860 0.016880

Connection atlas.toMTG r (Middle Temporal Gyrus, temporooccipital part Right) - RedNucleus_L T(82) = 2.59 0.011437 0.017215

Connection atlas.pITG r (Inferior Temporal Gyrus, posterior division Right) - SubstantiaNigra_L T(82) = 2.62 0.010523 0.020063

Connection atlas.aITG l (Inferior Temporal Gyrus, anterior division Left) - VentralTegmentalArea T(82) = 2.49 0.014841 0.027925

Connection atlas.pITG l (Inferior Temporal Gyrus, posterior division Left) - DorsalRapheNucleus T(82) = 2.48 0.015095 0.028402

Connection atlas.aTFusC r (Temporal Fusiform Cortex, anterior division Right) - PeriaqueductalGray T(82) = 2.68 0.008824 0.029345

Connection atlas.TOFusC l (Temporal Occipital Fusiform Cortex Left) - RedNucleus_R T(82) = 2.35 0.021097 0.032792

Connection atlas.pTFusC r (Temporal Fusiform Cortex, posterior division Right) - PeriaqueductalGray T(82) = 2.37 0.020068 0.034730

Connection atlas.sLOC l (Lateral Occipital Cortex, superior division Left) - RedNucleus_R T(82) = 2.27 0.025796 0.036165

Connection atlas.iLOC l (Lateral Occipital Cortex, inferior division Left) - VentralTegmentalArea T(82) = -2.32 0.022706 0.036897

Connection atlas.aITG l (Inferior Temporal Gyrus, anterior division Left) - SubThalamicNucleus_R T(82) = 2.37 0.020249 0.037605

Connection atlas.OFusG l (Occipital Fusiform Gyrus Left) - VentralTegmentalArea T(82) = -2.28 0.024909 0.044451

Connection atlas.OFusG l (Occipital Fusiform Gyrus Left) - SubstantiaNigra_L T(82) = 2.27 0.025741 0.044889

Connection atlas.pITG r (Inferior Temporal Gyrus, posterior division Right) - RedNucleus_L T(82) = 2.28 0.025093 0.045422

Connection atlas.OFusG r (Occipital Fusiform Gyrus Right) - VentralTegmentalArea T(82) = -2.27 0.025714 0.045964

Connection atlas.aITG l (Inferior Temporal Gyrus, anterior division Left) - LocusCoeruleus_L T(82) = 2.27 0.025806 0.047312

Connection atlas.pTFusC l (Temporal Fusiform Cortex, posterior division Left) - PeriaqueductalGray T(82) = 2.13 0.036065 0.050562

Connection atlas.pTFusC r (Temporal Fusiform Cortex, posterior division Right) - LocusCoeruleus_R T(82) = 2.18 0.032250 0.051817

Connection atlas.aTFusC r (Temporal Fusiform Cortex, anterior division Right) - VentralTegmentalArea T(82) = -2.41 0.018351 0.053554

Connection atlas.toITG r (Inferior Temporal Gyrus, temporooccipital part Right) - RedNucleus_L T(82) = 2.20 0.030950 0.053973

Connection atlas.sLOC l (Lateral Occipital Cortex, superior division Left) - SubstantiaNigra_R T(82) = 2.09 0.039852 0.054796

Connection atlas.TOFusC l (Temporal Occipital Fusiform Cortex Left) - SubstantiaNigra_L T(82) = 2.09 0.039966 0.058558

Connection atlas.toMTG l (Middle Temporal Gyrus, temporooccipital part Left) - PeriaqueductalGray T(82) = 2.08 0.040519 0.059125

Connection atlas.toMTG r (Middle Temporal Gyrus, temporooccipital part Right) - PeriaqueductalGray T(82) = 2.04 0.044749 0.062737

Connection atlas.toMTG r (Middle Temporal Gyrus, temporooccipital part Right) - NucleusTractusSolitarius_L T(82) = -2.00 0.048615 0.067494

Connection atlas.aITG l (Inferior Temporal Gyrus, anterior division Left) - SubThalamicNucleus_L T(82) = -1.99 0.049682 0.086641

Connection atlas.aTFusC r (Temporal Fusiform Cortex, anterior division Right) - LocusCoeruleus_R T(82) = 2.09 0.039994 0.100337

Connection atlas.aTFusC l (Temporal Fusiform Cortex, anterior division Left) - SubThalamicNucleus_R T(82) = 2.05 0.043977 0.103094

Cluster 89/120 F(3,80) = 29.48 0.000000 0.000000

Connection atlas.Caudate r - VentralTegmentalArea T(82) = 5.50 0.000000 0.000002

Connection atlas.Caudate r - RedNucleus_R T(82) = 4.86 0.000005 0.000016

Connection atlas.Pallidum r - PeriaqueductalGray T(82) = 5.05 0.000003 0.000029

Connection atlas.Pallidum r - RedNucleus_R T(82) = 4.95 0.000004 0.000034

Connection atlas.Putamen r - VentralTegmentalArea T(82) = 4.44 0.000028 0.000073

Connection atlas.Putamen l - VentralTegmentalArea T(82) = 4.41 0.000031 0.000082

Connection atlas.Caudate l - RedNucleus_L T(82) = 4.38 0.000035 0.000120

Connection atlas.Putamen l - RedNucleus_R T(82) = 4.28 0.000051 0.000128

Connection atlas.Caudate l - RedNucleus_R T(82) = 4.31 0.000044 0.000148

Connection atlas.Putamen r - RedNucleus_R T(82) = 4.21 0.000065 0.000166

Connection atlas.Caudate l - VentralTegmentalArea T(82) = 3.98 0.000147 0.000438

Connection atlas.Putamen r - RedNucleus_L T(82) = 3.67 0.000429 0.000945

Connection atlas.Pallidum l - VentralTegmentalArea T(82) = 3.91 0.000190 0.000971

Connection atlas.Putamen l - SubstantiaNigra_R T(82) = 3.61 0.000525 0.001155

Connection atlas.Pallidum r - VentralTegmentalArea T(82) = 3.64 0.000482 0.001814

Connection atlas.Pallidum l - PeriaqueductalGray T(82) = 3.56 0.000615 0.002587

Connection atlas.Caudate r - SubstantiaNigra_R T(82) = 2.93 0.004332 0.007554

Connection atlas.Caudate r - RedNucleus_L T(82) = 2.88 0.005052 0.008600

Connection atlas.Pallidum l - RedNucleus_R T(82) = 3.03 0.003308 0.009854

Connection atlas.Pallidum r - SubstantiaNigra_R T(82) = 2.81 0.006245 0.015855

Connection atlas.Putamen l - SubstantiaNigra_L T(82) = 2.46 0.015822 0.026618

Connection atlas.Pallidum l - SubstantiaNigra_L T(82) = 2.51 0.014114 0.031537

Connection atlas.Putamen l - DorsalRapheNucleus T(82) = -2.28 0.024989 0.040608

Connection atlas.Putamen r - DorsalRapheNucleus T(82) = -2.25 0.027422 0.043092

Connection atlas.Putamen r - LocusCoeruleus_L T(82) = 2.05 0.043413 0.066044

Cluster 92/120 F(3,80) = 26.95 0.000000 0.000000

Connection atlas.PaCiG l (Paracingulate Gyrus Left) - VentralTegmentalArea T(82) = 6.54 0.000000 0.000000

Connection atlas.PaCiG r (Paracingulate Gyrus Right) - VentralTegmentalArea T(82) = 6.10 0.000000 0.000000

Connection atlas.AC (Cingulate Gyrus, anterior division) - VentralTegmentalArea T(82) = 4.55 0.000018 0.000036

Connection atlas.IC l (Insular Cortex Left) - RedNucleus_R T(82) = 4.27 0.000051 0.000098

Connection atlas.PaCiG l (Paracingulate Gyrus Left) - RedNucleus_R T(82) = 4.29 0.000049 0.000098

Connection atlas.PaCiG l (Paracingulate Gyrus Left) - SubstantiaNigra_R T(82) = 4.27 0.000052 0.000103

Connection atlas.IC l (Insular Cortex Left) - PeriaqueductalGray T(82) = 4.11 0.000094 0.000173

Connection atlas.IC r (Insular Cortex Right) - RedNucleus_L T(82) = 3.91 0.000190 0.000335

Connection atlas.IC r (Insular Cortex Right) - RedNucleus_R T(82) = 3.89 0.000201 0.000350

Connection atlas.IC r (Insular Cortex Right) - DorsalRapheNucleus T(82) = -3.84 0.000242 0.000412

Connection atlas.IC l (Insular Cortex Left) - VentralTegmentalArea T(82) = 3.71 0.000372 0.000656

Connection atlas.PaCiG r (Paracingulate Gyrus Right) - RedNucleus_R T(82) = 3.69 0.000397 0.000747

Connection atlas.IC l (Insular Cortex Left) - RedNucleus_L T(82) = 3.66 0.000443 0.000772

Connection atlas.AC (Cingulate Gyrus, anterior division) - RedNucleus_R T(82) = 3.40 0.001044 0.001843

Connection atlas.IC l (Insular Cortex Left) - SubstantiaNigra_L T(82) = 3.31 0.001400 0.002301

Connection atlas.IC l (Insular Cortex Left) - DorsalRapheNucleus T(82) = -3.22 0.001858 0.002953

Connection atlas.pSMG r (Supramarginal Gyrus, posterior division Right) - RedNucleus_L T(82) = 3.22 0.001821 0.003175

Connection atlas.pSMG r (Supramarginal Gyrus, posterior division Right) - DorsalRapheNucleus T(82) = -3.13 0.002434 0.004095

Connection atlas.PaCiG r (Paracingulate Gyrus Right) - SubstantiaNigra_R T(82) = 3.11 0.002588 0.004158

Connection atlas.pSMG l (Supramarginal Gyrus, posterior division Left) - DorsalRapheNucleus T(82) = -3.06 0.002994 0.005560

Connection atlas.PaCiG r (Paracingulate Gyrus Right) - SubstantiaNigra_L T(82) = 2.96 0.003969 0.006102

Connection atlas.IC r (Insular Cortex Right) - PeriaqueductalGray T(82) = 2.94 0.004263 0.006350

Connection atlas.FO l (Frontal Operculum Cortex Left) - RedNucleus_R T(82) = 2.86 0.005311 0.008534

Connection atlas.AC (Cingulate Gyrus, anterior division) - LocusCoeruleus_L T(82) = 2.74 0.007571 0.011897

Connection atlas.pSMG r (Supramarginal Gyrus, posterior division Right) - SubstantiaNigra_L T(82) = 2.70 0.008532 0.013262

Connection atlas.PaCiG r (Paracingulate Gyrus Right) - RedNucleus_L T(82) = 2.62 0.010574 0.015583

Connection atlas.AC (Cingulate Gyrus, anterior division) - SubstantiaNigra_R T(82) = 2.62 0.010446 0.016236

Connection atlas.PaCiG l (Paracingulate Gyrus Left) - SubThalamicNucleus_R T(82) = 2.57 0.011901 0.017191

Connection atlas.IC r (Insular Cortex Right) - VentralTegmentalArea T(82) = 2.37 0.019893 0.027353

Connection atlas.PaCiG r (Paracingulate Gyrus Right) - LocusCoeruleus_L T(82) = 2.36 0.020771 0.028838

Connection atlas.AC (Cingulate Gyrus, anterior division) - SubstantiaNigra_L T(82) = 2.35 0.021119 0.031789

Connection atlas.pSMG l (Supramarginal Gyrus, posterior division Left) - SubstantiaNigra_L T(82) = 2.39 0.019350 0.032554

Connection atlas.pSMG r (Supramarginal Gyrus, posterior division Right) - NucleusTractusSolitarius_R T(82) = 2.27 0.026123 0.037355

Connection atlas.FO r (Frontal Operculum Cortex Right) - VentralTegmentalArea T(82) = 2.24 0.027969 0.038458

Connection atlas.IC r (Insular Cortex Right) - SubstantiaNigra_L T(82) = 2.21 0.030040 0.040147

Connection atlas.FO r (Frontal Operculum Cortex Right) - SubstantiaNigra_L T(82) = 2.20 0.030287 0.041248

Connection atlas.AC (Cingulate Gyrus, anterior division) - PeriaqueductalGray T(82) = 2.21 0.029825 0.043520

Connection atlas.pSMG l (Supramarginal Gyrus, posterior division Left) - RedNucleus_R T(82) = 2.18 0.032006 0.050854

Connection atlas.FO r (Frontal Operculum Cortex Right) - RedNucleus_L T(82) = 2.09 0.039369 0.052614

Connection atlas.FO r (Frontal Operculum Cortex Right) - LocusCoeruleus_L T(82) = 2.04 0.044650 0.058372

Cluster 99/120 F(3,80) = 20.82 0.000000 0.000000

Connection atlas.Accumbens r - VentralTegmentalArea T(82) = 6.55 0.000000 0.000000

Connection atlas.Accumbens l - VentralTegmentalArea T(82) = 5.63 0.000000 0.000001

Connection atlas.Accumbens r - RedNucleus_R T(82) = 4.89 0.000005 0.000023

Connection atlas.Accumbens r - RedNucleus_L T(82) = 4.22 0.000064 0.000261

Connection atlas.Amygdala l - DorsalRapheNucleus T(82) = 3.91 0.000188 0.000389

Connection atlas.FOrb r (Frontal Orbital Cortex Right) - RedNucleus_L T(82) = 3.81 0.000271 0.000530

Connection atlas.Accumbens l - SubstantiaNigra_R T(82) = 3.94 0.000169 0.000588

Connection atlas.Accumbens r - SubstantiaNigra_R T(82) = 3.89 0.000201 0.000757

Connection atlas.Accumbens l - RedNucleus_L T(82) = 3.74 0.000345 0.001007

Connection atlas.FOrb l (Frontal Orbital Cortex Left) - VentralTegmentalArea T(82) = 3.53 0.000679 0.001214

Connection atlas.FOrb l (Frontal Orbital Cortex Left) - RedNucleus_R T(82) = 3.50 0.000766 0.001352

Connection atlas.Amygdala r - VentralTegmentalArea T(82) = 3.07 0.002864 0.005316

Connection atlas.FOrb r (Frontal Orbital Cortex Right) - SubstantiaNigra_L T(82) = 3.02 0.003377 0.005427

Connection atlas.Amygdala l - LocusCoeruleus_R T(82) = 3.06 0.002977 0.005529

Connection atlas.Amygdala r - DorsalRapheNucleus T(82) = 2.80 0.006402 0.011029

Connection atlas.FOrb r (Frontal Orbital Cortex Right) - SubstantiaNigra_R T(82) = 2.75 0.007338 0.011406

Connection atlas.Accumbens l - RedNucleus_R T(82) = 2.77 0.006867 0.017227

Connection atlas.Amygdala l - LocusCoeruleus_L T(82) = 2.63 0.010121 0.017438

Connection atlas.FOrb r (Frontal Orbital Cortex Right) - VentralTegmentalArea T(82) = 2.58 0.011546 0.017753

Connection atlas.FOrb l (Frontal Orbital Cortex Left) - SubstantiaNigra_R T(82) = 2.44 0.017025 0.025098

Connection atlas.Amygdala r - LocusCoeruleus_L T(82) = 2.35 0.021136 0.034346

Connection atlas.FOrb r (Frontal Orbital Cortex Right) - LocusCoeruleus_L T(82) = 2.28 0.025454 0.036399

Connection atlas.FOrb r (Frontal Orbital Cortex Right) - RedNucleus_R T(82) = 2.17 0.033105 0.046412

Cluster 101/120 F(3,80) = 16.92 0.000000 0.000000

Connection atlas.SFG l (Superior Frontal Gyrus Left) - RedNucleus_R T(82) = 5.42 0.000001 0.000001

Connection atlas.SFG l (Superior Frontal Gyrus Left) - VentralTegmentalArea T(82) = 3.88 0.000210 0.000412

Connection atlas.FP r (Frontal Pole Right) - RedNucleus_L T(82) = 3.86 0.000225 0.000494

Connection atlas.FP r (Frontal Pole Right) - SubstantiaNigra_L T(82) = 3.77 0.000305 0.000661

Connection atlas.FP l (Frontal Pole Left) - RedNucleus_R T(82) = 3.70 0.000387 0.000712

Connection atlas.SFG r (Superior Frontal Gyrus Right) - VentralTegmentalArea T(82) = 3.47 0.000821 0.001726

Connection atlas.FP l (Frontal Pole Left) - NucleusTractusSolitarius_R T(82) = -3.28 0.001505 0.002474

Connection atlas.SFG r (Superior Frontal Gyrus Right) - RedNucleus_R T(82) = 3.29 0.001465 0.002832

Connection atlas.SFG l (Superior Frontal Gyrus Left) - SubstantiaNigra_R T(82) = 3.03 0.003251 0.005669

Connection atlas.FP r (Frontal Pole Right) - LocusCoeruleus_R T(82) = -2.95 0.004144 0.007315

Connection atlas.SFG r (Superior Frontal Gyrus Right) - SubstantiaNigra_R T(82) = 2.93 0.004406 0.007976

Connection atlas.SFG r (Superior Frontal Gyrus Right) - RedNucleus_L T(82) = 2.92 0.004540 0.008116

Connection atlas.FP l (Frontal Pole Left) - VentralTegmentalArea T(82) = 2.81 0.006152 0.009359

Connection atlas.FP l (Frontal Pole Left) - DorsalRapheNucleus T(82) = -2.49 0.014753 0.021310

Connection atlas.SFG r (Superior Frontal Gyrus Right) - SubstantiaNigra_L T(82) = 2.46 0.016001 0.025144

Connection atlas.SFG r (Superior Frontal Gyrus Right) - DorsalRapheNucleus T(82) = -2.20 0.030374 0.046207

Connection atlas.FP r (Frontal Pole Right) - VentralTegmentalArea T(82) = 2.19 0.031191 0.050116

Connection atlas.FP r (Frontal Pole Right) - DorsalRapheNucleus T(82) = -2.11 0.037948 0.059632

Connection atlas.FP r (Frontal Pole Right) - NucleusTractusSolitarius_L T(82) = -2.10 0.038768 0.060260

Connection atlas.FP r (Frontal Pole Right) - RedNucleus_R T(82) = 2.00 0.049068 0.073860

Cluster 104/120 F(3,80) = 15.63 0.000000 0.000000

Connection atlas.TP l (Temporal Pole Left) - DorsalRapheNucleus T(82) = 5.37 0.000001 0.000002

Connection atlas.AG r (Angular Gyrus Right) - RedNucleus_L T(82) = 4.80 0.000007 0.000018

Connection atlas.TP r (Temporal Pole Right) - DorsalRapheNucleus T(82) = 4.42 0.000030 0.000055

Connection atlas.TP l (Temporal Pole Left) - LocusCoeruleus_L T(82) = 4.17 0.000074 0.000141

Connection atlas.IFG oper l (Inferior Frontal Gyrus, pars opercularis Left) - RedNucleus_R T(82) = 4.06 0.000110 0.000334

Connection atlas.IFG tri l (Inferior Frontal Gyrus, pars triangularis Left) - RedNucleus_R T(82) = 3.87 0.000215 0.000698

Connection atlas.IFG oper r (Inferior Frontal Gyrus, pars opercularis Right) - DorsalRapheNucleus T(82) = -3.66 0.000452 0.000808

Connection atlas.TP r (Temporal Pole Right) - LocusCoeruleus_L T(82) = 3.63 0.000499 0.000811

Connection atlas.pMTG r (Middle Temporal Gyrus, posterior division Right) - NucleusTractusSolitarius_L T(82) = -3.58 0.000572 0.001259

Connection atlas.aSMG l (Supramarginal Gyrus, anterior division Left) - RedNucleus_R T(82) = 3.37 0.001154 0.002063

Connection atlas.aSMG r (Supramarginal Gyrus, anterior division Right) - DorsalRapheNucleus T(82) = -3.05 0.003056 0.004967

Connection atlas.IFG tri r (Inferior Frontal Gyrus, pars triangularis Right) - DorsalRapheNucleus T(82) = -3.05 0.003121 0.005649

Connection atlas.aMTG r (Middle Temporal Gyrus, anterior division Right) - VentralTegmentalArea T(82) = 2.99 0.003643 0.006352

Connection atlas.aMTG r (Middle Temporal Gyrus, anterior division Right) - SubstantiaNigra_R T(82) = 2.96 0.004071 0.006931

Connection atlas.aSMG r (Supramarginal Gyrus, anterior division Right) - NucleusTractusSolitarius_R T(82) = 2.86 0.005443 0.008553

Connection atlas.pMTG r (Middle Temporal Gyrus, posterior division Right) - NucleusTractusSolitarius_R T(82) = -2.89 0.004899 0.008869

Connection atlas.AG l (Angular Gyrus Left) - DorsalRapheNucleus T(82) = -2.77 0.006860 0.010436

Connection atlas.IFG tri l (Inferior Frontal Gyrus, pars triangularis Left) - DorsalRapheNucleus T(82) = -2.86 0.005381 0.012024

Connection atlas.AG r (Angular Gyrus Right) - SubstantiaNigra_L T(82) = 2.75 0.007286 0.012953

Connection atlas.pMTG l (Middle Temporal Gyrus, posterior division Left) - NucleusTractusSolitarius_R T(82) = -2.72 0.007896 0.013129

Connection atlas.IFG tri r (Inferior Frontal Gyrus, pars triangularis Right) - RedNucleus_L T(82) = 2.65 0.009641 0.016031

Connection atlas.IFG oper r (Inferior Frontal Gyrus, pars opercularis Right) - RedNucleus_L T(82) = 2.62 0.010485 0.016477

Connection atlas.aMTG r (Middle Temporal Gyrus, anterior division Right) - SubThalamicNucleus_R T(82) = 2.59 0.011439 0.017976

Connection atlas.aSMG l (Supramarginal Gyrus, anterior division Left) - DorsalRapheNucleus T(82) = -2.58 0.011512 0.018090

Connection atlas.IFG oper r (Inferior Frontal Gyrus, pars opercularis Right) - SubstantiaNigra_L T(82) = 2.52 0.013595 0.020463

Connection atlas.IFG tri r (Inferior Frontal Gyrus, pars triangularis Right) - NucleusTractusSolitarius_L T(82) = -2.53 0.013488 0.021431

Connection atlas.AG r (Angular Gyrus Right) - SubstantiaNigra_R T(82) = 2.52 0.013596 0.022608

Connection atlas.TP l (Temporal Pole Left) - LocusCoeruleus_R T(82) = 2.40 0.018441 0.025115

Connection atlas.IFG oper l (Inferior Frontal Gyrus, pars opercularis Left) - VentralTegmentalArea T(82) = 2.50 0.014258 0.026479

Connection atlas.TP r (Temporal Pole Right) - SubstantiaNigra_L T(82) = 2.36 0.020762 0.029395

Connection atlas.aSMG r (Supramarginal Gyrus, anterior division Right) - VentralTegmentalArea T(82) = -2.35 0.021236 0.030987

Connection atlas.IFG oper r (Inferior Frontal Gyrus, pars opercularis Right) - RedNucleus_R T(82) = 2.25 0.027152 0.037333

Connection atlas.pMTG r (Middle Temporal Gyrus, posterior division Right) - LocusCoeruleus_R T(82) = -2.31 0.023454 0.037685

Connection atlas.pMTG r (Middle Temporal Gyrus, posterior division Right) - DorsalRapheNucleus T(82) = -2.22 0.029382 0.046685

Connection atlas.AG r (Angular Gyrus Right) - DorsalRapheNucleus T(82) = -2.17 0.032850 0.049448

Connection atlas.IFG tri r (Inferior Frontal Gyrus, pars triangularis Right) - RedNucleus_R T(82) = 2.11 0.037920 0.054773

Connection atlas.AG l (Angular Gyrus Left) - SubstantiaNigra_R T(82) = 2.08 0.040505 0.058507

Connection atlas.aSMG l (Supramarginal Gyrus, anterior division Left) - NucleusTractusSolitarius_R T(82) = 2.08 0.040650 0.059316

Connection atlas.IFG tri r (Inferior Frontal Gyrus, pars triangularis Right) - SubstantiaNigra_L T(82) = 2.06 0.042752 0.060369

Connection atlas.AG l (Angular Gyrus Left) -RedNucleus_R T(82) = 2.01 0.047361 0.065754

Connection atlas.aSMG l (Supramarginal Gyrus, anterior division Left) - SubThalamicNucleus_R T(82) = -2.01 0.047399 0.067781

Connection atlas.pMTG l (Middle Temporal Gyrus, posterior division Left) - VentralTegmentalArea T(82) = 2.00 0.048483 0.074549

Connection atlas.aSTG l (Superior Temporal Gyrus, anterior division Left) - SubstantiaNigra_L T(82) = 2.00 0.049296 0.076623

Cluster 108/120 F(3,80) = 12.97 0.000001 0.000001

Connection atlas.MedFC (Frontal Medial Cortex) - VentralTegmentalArea T(82) = 4.89 0.000005 0.000012

Connection atlas.PC (Cingulate Gyrus, posterior division) - VentralTegmentalArea T(82) = 4.38 0.000035 0.000063

Connection atlas.PC (Cingulate Gyrus, posterior division) - SubstantiaNigra_R T(82) = 3.62 0.000512 0.000814

Connection atlas.MedFC (Frontal Medial Cortex) - SubstantiaNigra_R T(82) = 3.54 0.000666 0.001253

Connection atlas.MedFC (Frontal Medial Cortex) - RedNucleus_R T(82) = 3.26 0.001635 0.002923

Connection atlas.PC (Cingulate Gyrus, posterior division) - LocusCoeruleus_L T(82) = 3.18 0.002081 0.003166

Connection atlas.Precuneous (Precuneous Cortex) - LocusCoeruleus_L T(82) = 2.96 0.004075 0.007471

Connection atlas.Precuneous (Precuneous Cortex) - VentralTegmentalArea T(82) = 2.60 0.010996 0.018944

Connection atlas.PC (Cingulate Gyrus, posterior division) - RedNucleus_R T(82) = 2.48 0.015228 0.021560

Connection atlas.Precuneous (Precuneous Cortex) - SubstantiaNigra_R T(82) = 2.52 0.013821 0.023206

Connection atlas.PC (Cingulate Gyrus, posterior division) - SubstantiaNigra_L T(82) = 2.43 0.017295 0.024247

Connection atlas.SubCalC (Subcallosal Cortex) - DorsalRapheNucleus T(82) = 2.49 0.014681 0.025293

Connection atlas.SubCalC (Subcallosal Cortex) - SubstantiaNigra_R T(82) = 2.39 0.019363 0.032196

Connection atlas.SubCalC (Subcallosal Cortex) - SubThalamicNucleus_R T(82) = 2.36 0.020716 0.033794

Connection atlas.PC (Cingulate Gyrus, posterior division) - SubThalamicNucleus_R T(82) = 2.27 0.026029 0.035449

Connection atlas.SubCalC (Subcallosal Cortex) - VentralTegmentalArea T(82) = 2.30 0.024031 0.038182

Connection atlas.Precuneous (Precuneous Cortex) - SubstantiaNigra_L T(82) = 2.24 0.027757 0.042227

Connection atlas.SubCalC (Subcallosal Cortex) - RedNucleus_L T(82) = 2.18 0.031854 0.048980

Connection atlas.MedFC (Frontal Medial Cortex) - SubThalamicNucleus_R T(82) = 2.06 0.042942 0.061407

Cluster 112/120 F(3,80) = 7.11 0.000269 0.000288

Connection atlas.MidFG l (Middle Frontal Gyrus Left) - RedNucleus_R T(82) = 4.84 0.000006 0.000015

Connection atlas.MidFG r (Middle Frontal Gyrus Right) - RedNucleus_L T(82) = 3.65 0.000466 0.001058

Connection atlas.SPL r (Superior Parietal Lobule Right) - NucleusTractusSolitarius_R T(82) = 2.80 0.006401 0.010463

Connection atlas.SPL l (Superior Parietal Lobule Left) - VentralTegmentalArea T(82) = -2.58 0.011738 0.019074

Connection atlas.MidFG r (Middle Frontal Gyrus Right) - SubstantiaNigra_L T(82) = 2.53 0.013481 0.023227

Connection atlas.SPL r (Superior Parietal Lobule Right) - VentralTegmentalArea T(82) = -2.48 0.014998 0.023312

Connection atlas.MidFG l (Middle Frontal Gyrus Left) - LocusCoeruleus_R T(82) = 2.40 0.018531 0.028190

Connection atlas.MidFG l (Middle Frontal Gyrus Left) - VentralTegmentalArea T(82) = 2.39 0.019009 0.028614

Connection atlas.SPL r (Superior Parietal Lobule Right) - DorsalRapheNucleus T(82) = -2.37 0.020111 0.030923

Connection atlas.MidFG r (Middle Frontal Gyrus Right) - RedNucleus_R T(82) = 2.36 0.020577 0.033823

Cluster 114/120 F(3,80) = 5.80 0.001226 0.001290

Connection atlas.HG l (Heschl's Gyrus Left) - RedNucleus_R T(82) = 3.98 0.000147 0.000305

Connection atlas.HG r (Heschl's Gyrus Right) - RedNucleus_R T(82) = 3.77 0.000303 0.000578

Connection atlas.HG r (Heschl's Gyrus Right) - VentralTegmentalArea T(82) = 3.32 0.001332 0.002326

Connection atlas.HG r (Heschl's Gyrus Right) - DorsalRapheNucleus T(82) = -3.30 0.001426 0.002457

Connection atlas.HG l (Heschl's Gyrus Left) - VentralTegmentalArea T(82) = 3.24 0.001726 0.003205

Connection atlas.HG l (Heschl's Gyrus Left) - SubstantiaNigra_L T(82) = 3.05 0.003113 0.005564

Connection atlas.pSTG r (Superior Temporal Gyrus, posterior division Right) - DorsalRapheNucleus T(82) = -2.84 0.005645 0.009845

Connection atlas.HG r (Heschl's Gyrus Right) - PeriaqueductalGray T(82) = 2.64 0.009895 0.015988

Connection atlas.HG r (Heschl's Gyrus Right) - SubstantiaNigra_L T(82) = 2.52 0.013671 0.021484

Connection atlas.PP l (Planum Polare Left) -VentralTegmentalArea T(82) = -2.41 0.018288 0.029718

Connection atlas.PT l (Planum Temporale Left) - SubstantiaNigra_L T(82) = 2.18 0.032211 0.048485

Connection atlas.PP l (Planum Polare Left) - LocusCoeruleus_L T(82) = 2.18 0.032482 0.049415

Connection atlas.HG l (Heschl's Gyrus Left) - PeriaqueductalGray T(82) = 2.06 0.042536 0.062707

Connection atlas.HG r (Heschl's Gyrus Right) - SubstantiaNigra_R T(82) = 2.04 0.044208 0.065173

Connection atlas.PP l (Planum Polare Left) - RedNucleus_L T(82) = 2.01 0.048141 0.070246

Cluster 118/120 F(3,80) = 2.89 0.040513 0.041200

Connection atlas.LG l (Lingual Gyrus Left) - PeriaqueductalGray T(82) = 3.55 0.000647 0.001251

Connection atlas.LG l (Lingual Gyrus Left) - LocusCoeruleus_L T(82) = 3.32 0.001346 0.002500

Connection atlas.LG r (Lingual Gyrus Right) - PeriaqueductalGray T(82) = 3.10 0.002619 0.005061

Connection atlas.ICC l (Intracalcarine Cortex Left) - PeriaqueductalGray T(82) = 3.04 0.003132 0.006370

Connection atlas.LG r (Lingual Gyrus Right) - SubstantiaNigra_R T(82) = 2.93 0.004327 0.008036

Connection atlas.ICC r (Intracalcarine Cortex Right) - PeriaqueductalGray T(82) = 2.83 0.005805 0.011068

Connection atlas.SCC r (Supracalcarine Cortex Right) - SubstantiaNigra_L T(82) = 2.85 0.005469 0.012821

Connection atlas.LG r (Lingual Gyrus Right) - SubstantiaNigra_L T(82) = 2.63 0.010297 0.017740

Connection atlas.Cuneal l (Cuneal Cortex Left) - LocusCoeruleus_R T(82) = -2.68 0.008818 0.017761

Connection atlas.ICC r (Intracalcarine Cortex Right) - SubstantiaNigra_L T(82) = 2.60 0.010948 0.018862

Connection atlas.ICC l (Intracalcarine Cortex Left) - SubstantiaNigra_L T(82) = 2.60 0.011005 0.019428

Connection atlas.LG r (Lingual Gyrus Right) - LocusCoeruleus_L T(82) = 2.58 0.011621 0.019784

Connection atlas.LG l (Lingual Gyrus Left) - VentralTegmentalArea T(82) = 2.38 0.019727 0.031696

Connection atlas.ICC r (Intracalcarine Cortex Right) - SubstantiaNigra_R T(82) = 2.31 0.023527 0.039120

Connection atlas.ICC r (Intracalcarine Cortex Right) - LocusCoeruleus_L T(82) = 2.28 0.025458 0.041845

Connection atlas.Cuneal l (Cuneal Cortex Left) - SubstantiaNigra_L T(82) = 2.20 0.030577 0.050842

Connection atlas.LG l (Lingual Gyrus Left) - SubstantiaNigra_L T(82) = 2.14 0.035589 0.054140

Connection atlas.LG l (Lingual Gyrus Left) - DorsalRapheNucleus T(82) = 2.06 0.042307 0.062076

Connection atlas.LG l (Lingual Gyrus Left) - SubstantiaNigra_R T(82) = 2.04 0.044461 0.064221

Connection atlas.Cuneal r (Cuneal Cortex Right) - RedNucleus_R T(82) = -2.13 0.035865 0.064919

Connection atlas.LG r (Lingual Gyrus Right) - VentralTegmentalArea T(82) = 2.02 0.046871 0.065073

Connection atlas.OP r (Occipital Pole Right) - VentralTegmentalArea T(82) = -2.12 0.037221 0.067375

Connection atlas.Cuneal r (Cuneal Cortex Right) - SubstantiaNigra_L T(82) = 2.10 0.039205 0.070079

Connection atlas.ICC l (Intracalcarine Cortex Left) - VentralTegmentalArea T(82) = 2.00 0.049316 0.078274

Connection atlas.ICC l (Intracalcarine Cortex Left) - SubstantiaNigra_R T(82) = 1.99 0.049811 0.078274

Connection atlas.OP r (Occipital Pole Right) - NucleusTractusSolitarius_R T(82) = -2.01 0.047464 0.082773

Cluster 119/120  F(3,80) = 2.78  0.046654  0.047046

Connection atlas.PreCG r (Precentral Gyrus Right) - DorsalRapheNucleus  T(82) = -3.49  0.000792 0.001364

Connection atlas.PostCG r (Postcentral Gyrus Right) - DorsalRapheNucleus  T(82) = -2.99  0.003724 0.007006

Connection atlas.PostCG r (Postcentral Gyrus Right) - RedNucleus_L  T(82) = -2.64  0.009872 0.017429

Connection atlas.PO l (Parietal Operculum Cortex Left) - PeriaqueductalGray  T(82) = 2.34  0.021837 0.033219

Connection atlas.PO r (Parietal Operculum Cortex Right) - DorsalRapheNucleus  T(82) = -2.29 0.024530  0.038546

Connection atlas.PostCG r (Postcentral Gyrus Right) - LocusCoeruleus_R  T(82) = -2.22  0.029347 0.045126

Connection atlas.PostCG l (Postcentral Gyrus Left) - RedNucleus_L  T(82) = -2.17  0.033034 0.055574

Connection atlas.CO l (Central Opercular Cortex Left) - PeriaqueductalGray  T(82) = 2.06  0.042285 0.062338

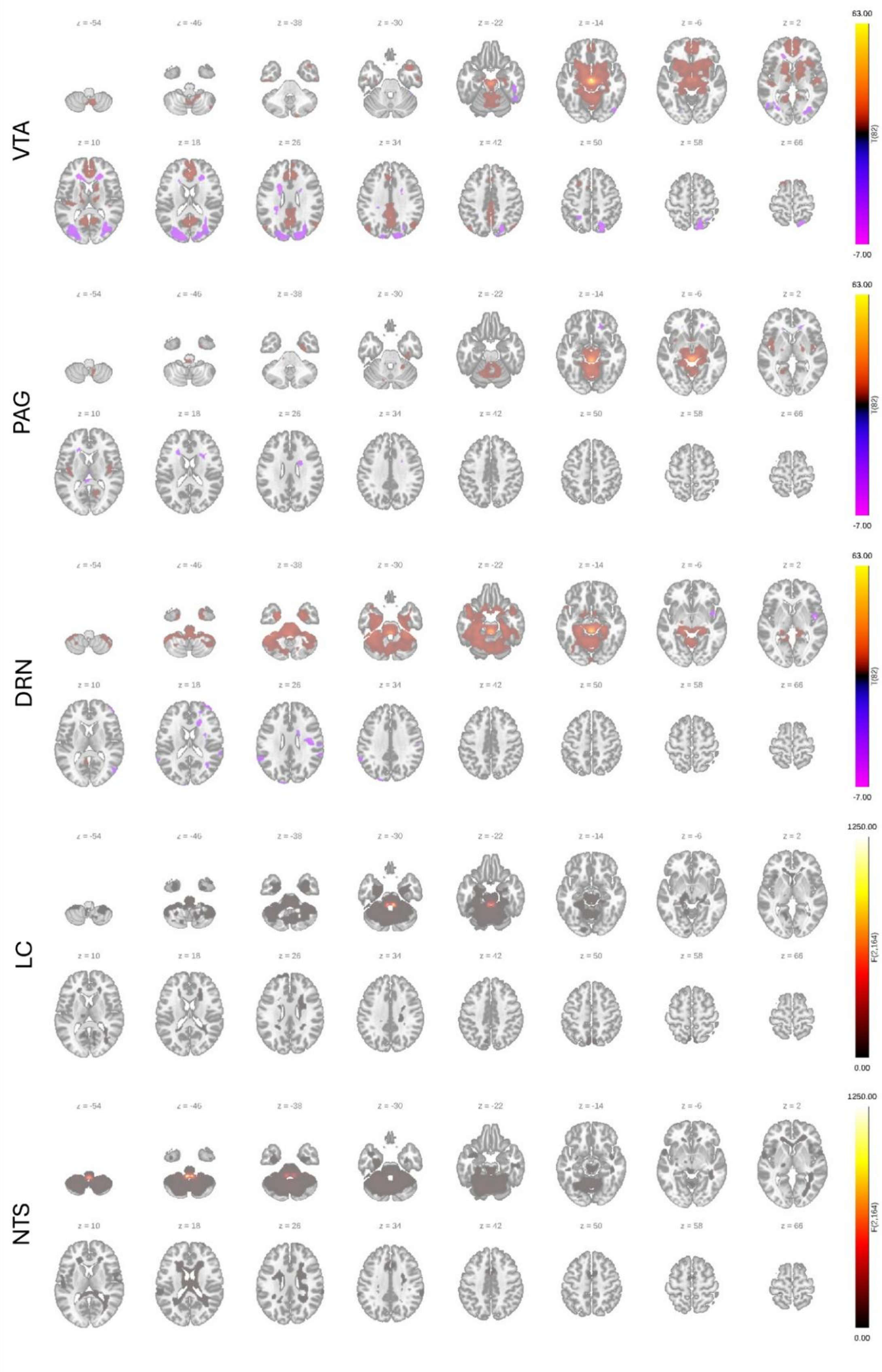


*Supplementary Figure 4. Baseline resting state brainstem functional connectivity of ventral tegmental area (VTA), periaqueductal gray (PAG), dorsal raphe nucleus (DRN), locus coeruleus (LC), nucleus tractus solitarius (NTS) assessed with seed-based connectivity (SBC). For bilateral seeds (LC, NTS) "any effect" was assessed.*

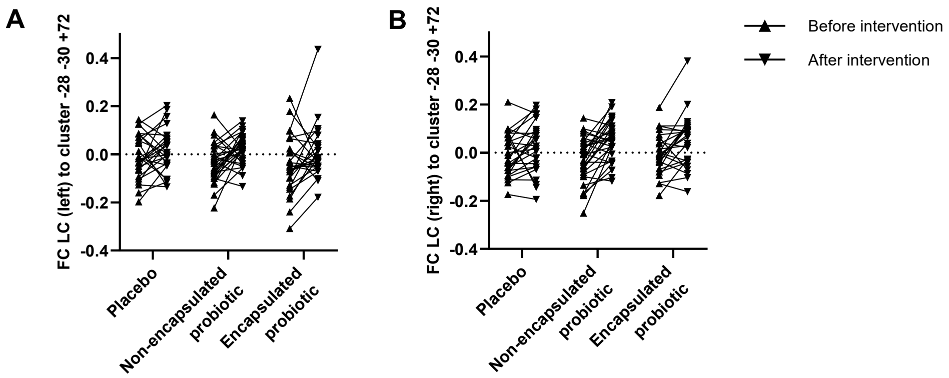


*Supplementary Figure 5. Brainstem-to-cortex functional connectivity at rest was affected by probiotic intervention healthy elderly. For the functional connectivity of both A left locus coeruleus (LC) and B right LC with a cluster covering left pre-, postcentral gyrus, superior parietal lobule a main time effect was observed. Upward triangles indicate baseline/before intervention per group, and downward triangles after intervention. n=83 participants. Seed-to-voxel analyses (SBC) performed in CONN Toolbox version 25.b. Figure created in GraphPad Prism.*

### *Supplementary References*


1. Rode J, Hutchinson AN, Chatzopoulou MS, et al. Micro-encapsulation differentially impacts probiotic effects on brain structure and function in an elderly population - A randomised placebo-controlled trial. *Brain Behav Immun* 2025;130:106113. doi: 10.1016/j.bbi.2025.106113 [published Online First: 20250919]
2. Whitfield-Gabrieli S, Nieto-Castanon A. Conn: a functional connectivity toolbox for correlated and anticorrelated brain networks. *Brain Connect* 2012;2(3):125-41. doi: 10.1089/brain.2012.0073 [published Online First: 2012/05/31]
3. Levinson S, Miller M, Iftekhar A, et al. A structural connectivity atlas of limbic brainstem nuclei. *Front Neuroimaging* 2022;1:1009399. doi: 10.3389/fnimg.2022.1009399 [published Online First: 20230112]
4. Xiao Y, Lau JC, Anderson T, et al. An accurate registration of the BigBrain dataset with the MNI PD25 and ICBM152 atlases. *Sci Data* 2019;6(1):210. doi: 10.1038/s41597-019-0217-0 [published Online First: 20191017]
